# Accelerated electrocardiographic aging of the heart as a risk determinant for atrial fibrillation: A Mendelian randomization study

**DOI:** 10.64898/2025.12.30.25343249

**Authors:** Seunghoon Cho, Seng Chan You, Taehyun Hwang, Hanjin Park, Daehoon Kim, Tae-Hoon Kim, Jae-Sun Uhm, Hui-Nam Pak, Pil-Sung Yang, Hee Tae Yu, Boyoung Joung

**Author notes:** Co-corresponding authors: **Hee Tae Yu, Associate Professor, MD, PhD,** 50-1 Yonsei-ro, Seodaemun-gu, Seoul 03722, Korea, Boyoung Joung, Professor, MD, PhD, 50-1 Yonsei-ro, Seodaemun-gu, Seoul 03722, Korea. Both authors contributed equally to this work (Seunghoon Cho, Seng Chan You).

## Abstract

**Background:** Artificial intelligence (AI)-derived electrocardiographic aging (ECG-aging) has been found to be associated with the risk of atrial fibrillation (AF).

**Objectives:** We aimed to assess the causal association between the discrepancy in AI-predicted electrocardiographic age and chronological age (AI-ECG age gap) and AF risk.

**Methods:** We analyzed 12-lead ECGs in UK Biobank to derive the AI-ECG age gap using our latest AI-based age prediction model. Associations between measured ECG-aging and genetically predicted ECG-aging based on a genetic risk score (GRS) were evaluated in relation to AF using multivariable regression and Mendelian randomization (MR). MR analyses incorporated GRS-based individual-level data and summary-level genome-wide association study (GWAS) data from large external consortia, with causal estimates obtained using inverse-variance weighting and complementary sensitivity methods. Mediation MR was additionally performed to assess intermediary causal pathways.

**Results:** Each 1-SD increase in the AI-ECG age gap was associated with an increased risk of AF in observational analyses (HR 1.43 [95% CI, 1.29–1.59]) and GRS-based MR (OR 1.06 [95% CI, 1.01–1.12]). Two-sample MR analyses in two large, independent GWAS datasets replicated these findings (OR 1.13 [95% CI, 1.02–1.25] in both), with multiple sensitivity analyses confirming robustness. Bidirectional MR suggested potential reverse causation from AF to ECG-aging. Non-linear MR demonstrated a consistent positive association across the AI-ECG age gap spectrum. Mediation MR further identified heart failure as a key intermediary, accounting for approximately 70% of the total causal effect of ECG-aging on AF.

**Conclusion:** This study provides genetic evidence supporting a causal association between ECG-aging and the risk of AF. These findings highlight ECG-aging as a causally relevant and clinically informative biomarker for AF that may help identify individuals at increased risk.

## Introduction

Electrocardiography (ECG) is a widely used, non-invasive tool recording the heart’s electrical activity and provides a cornerstone of cardiovascular health assessment.^1^ While chronological age is a major determinant of age-related cardiac conditions such as atrial fibrillation (AF), individuals of the same age often show considerable variability in disease susceptibility.^2,3^ This discrepancy may be better explained by physiological aging, which reflects organ-specific functional decline over time.^4^

Recent advances in artificial intelligence (AI) have enabled the estimation of physiological age from raw 12-lead ECG data.^5–7^ The difference between AI-predicted electrocardiographic age (AI-ECG age) and chronological age—termed the AI-ECG age gap or electrocardiographic aging (ECG-aging)—has been associated with cardiovascular health and adverse outcomes, including all-cause and cardiovascular mortality.^1,6,8,9^ Notably, individuals with a higher AI-ECG age than their chronological age appear to exhibit signs of accelerated ECG-aging and elevated cardiovascular risk.^8,10,11^ Building on this concept, our recent multinational study introduced and validated the PROPHECG-Age deep learning model, which was trained on over 1.5 million ECGs from diverse populations to predict AI-ECG age. We demonstrated that a larger AI-ECG age gap was robustly associated with both new-and early-onset AF, suggesting its potential as a novel biomarker for AF risk stratification.^7^ However, existing studies have been primarily observational, leaving uncertainty about whether ECG-aging causally contributes to AF due to possible residual confounding and reverse causation.

To overcome these limitations, we conducted a comprehensive Mendelian randomization (MR) analysis using data from the UK Biobank and large-scale external genome-wide association studies (GWAS). MR leverages genetic variants, typically single-nucleotide polymorphisms (SNPs), that are robustly associated with exposures to estimate their causal effects on outcomes of interest.^12,13^ Given the random allocation of alleles, MR mimics key features of randomized trials and mitigates concerns of residual confounding and reverse causation inherent to observational studies.^14^ Accordingly, we used genetic instruments for the AI-ECG age gap to assess its unconfounded, directional relationship with the risk of AF. Through this genetic approach, we aimed to determine whether ECG-aging serves not only as a prognostic correlate, but also as a potentially causal biomarker for AF risk.

## Methods

### Ethical consideration and approval

The UK Biobank received ethical approval from the North West Multi-Centre Research Ethics Committee (11/NW/0382). Comprehensive analyses and their derived data using the UK Biobank dataset were conducted under application number 77793. All study analyses were approved by the Institutional Review Board of the Yonsei University Health System (4-2025-1094). This study followed the Strengthening the Reporting of Observational Studies in Epidemiology using Mendelian Randomization (STROBE–MR) reporting guideline.

### Study population and participant selection

This study utilized data from the UK Biobank, a large-scale prospective cohort of over 500,000 individuals aged 40–69 years recruited between 2006 and 2010 across the UK, and the details of the data have been described elsewhere.^15,16^ After excluding participants without usable 12-lead ECG data, with first ECG acquisition after December 31, 2020, or of non–White British ancestry, a total of 36,044 participants with sufficient data to derive the AI-ECG age gap were included (**Figure 1**). Of these, 35,090 participants without prevalent AF were used in the observational analyses. For MR analyses, 29,806 participants passed genetic quality control, and 29,734 with complete individual-level genotype and phenotype data were included in the genetic risk score (GRS)-based MR. Summary-level two-sample MR analyses were also conducted using AF GWAS data from two large external consortia (N = 1,030,836 and N = 287,805).

**Figure 1.**
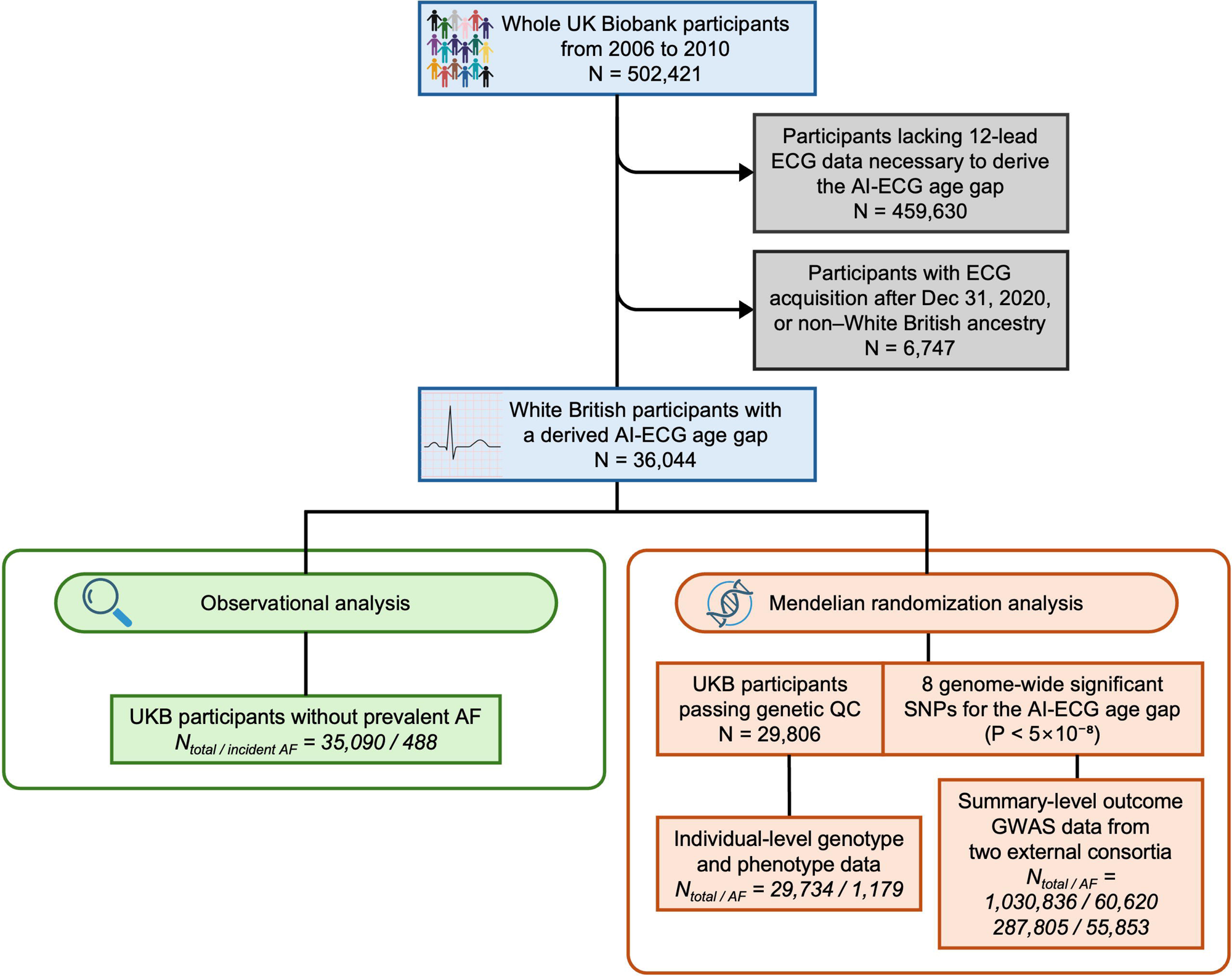
Flow diagram of sample selection, study design, and analysis. The AI-ECG age gap was derived using a fine-tuned version of the PROPHECG-Age model, as described in our previous publication.^7^ Eight genetic instruments reaching genome-wide significance for ECG-aging were obtained from the GWAS results reported by Shah et al.^21^ Abbreviations: AF, atrial fibrillation; AI, artificial intelligence; ECG, electrocardiogram; ECG-aging, electrocardiographic aging; GWAS, genome-wide association study; MR, Mendelian randomization; GRS, genetic risk score; QC, quality control; UKB, UK Biobank.

### Study variables and outcome assessment

In this study, we utilized a fine-tuned version of the PROPHECG-Age model to estimate AI-ECG age for UK Biobank participants. A detailed comparative results of the performance of this refined model, which was fine-tuned using a subset of the UK Biobank dataset, is provided in our previous publication.^7^ The AI-ECG age gap was calculated by subtracting each participant’s chronological age from their predicted AI-ECG age based on the first acquired ECG. For subsequent analyses, participants were stratified into quartiles of the AI-ECG age gap, with the highest quartile representing the largest positive age gap.

Baseline variables—including demographics, anthropometric data, comorbidities, lifestyle behaviors, and laboratory values—were collected based on the first ECG acquisition date (index date). Cardiac magnetic resonance (CMR) imaging data were obtained from the UK Biobank imaging study, which assessed cardiac structure and function using standardized acquisition protocols and quantification methods, as described elsewhere.^17,18^ The primary outcome was incident AF, defined as the first diagnosis occurring after the index date. Diagnoses were identified using the International Classification of Diseases, 10th Revision (ICD-10), and self-reported non-cancer illness codes, requiring either more than one inpatient or two outpatient records (or primary care records) to ensure accuracy. The AF definition based on the ICD-10 code has demonstrated a positive predictive value of 94.1% in our external validation study.^19^ A full list of diagnosis definitions and ICD-10 codes is available in **Supplemental Table S1**. Participants were followed until the first occurrence of AF, death, or December 31, 2020, whichever came first.

### Observational statistical analyses

Clinical characteristics were summarized as means with standard deviations (SD) for continuous variables and counts with percentages for categorical variables. Study group comparisons were performed using Pearson’s chi-squared or Fisher’s exact test for categorical variables, and either one-way ANOVA or the Kruskal–Wallis test for continuous variables, depending on distribution normality.

Adjusted event rates are presented as events per 1,000 person-years (PY), and cumulative incidence of incident AF across AI-ECG age gap quartiles was estimated using Cox proportional hazards models adjusted for age and sex, and visualized graphically. Hazard ratios (HR) with 95% confidence intervals (CI) were calculated from Cox models after confirming the proportional hazards assumption. To control for potential confounding, effect estimates from observational regression models were adjusted for chronological age, sex, Townsend deprivation index, smoking status, alcohol consumption, physical activity (PA) levels, body mass index (BMI), systolic blood pressure (BP), hemoglobin A1c, low-density lipoprotein (LDL) cholesterol, and estimated glomerular filtration rate (eGFR). As a sensitivity analysis, the Fine–Gray sub-distribution hazard model was applied to account for death as a competing risk.^20^

Statistical analyses were performed using R (version 4.2.3, The R Foundation; www.R-project.org), with two-sided p-values <0.05 considered statistically significant.

### Genetic instrumental variables, quality control, and genetic risk score derivation

Genetic instruments for the primary exposure, AI-ECG age gap, were selected from eight independent SNPs reaching genome-wide significance (P < 5×10^-8^) in a previous GWAS conducted in a subset of UK Biobank.^21^ For other traits evaluated as candidate mediators or outcomes, we used publicly available GWAS summary statistics from recent large consortia or meta-analyses to maximize sample size, ensure ancestral homogeneity, and minimize sample overlap with our primary dataset whenever possible. GWAS data sources for all phenotypes in this study are summarized in **Supplemental Table S2**.

Individual-level genotype data were obtained from UK Biobank Imputation V3 (GRCh37, BGEN format), and genetic quality control was performed following UK Biobank protocols.^15^ Participants were excluded if they had outlier heterozygosity or missingness, discordant genetic and reported sex, sex chromosome aneuploidy, or excess relatedness by kinship inference. A GRS for ECG-aging was then constructed by summing the product of GWAS-derived effect sizes (β coefficients) and allele dosages for each selected SNP using PLINK 2.0 (version alpha 6.9), yielding an individual-level measure of genetically predicted ECG-aging used as the exposure in subsequent GRS-based regression and MR analyses.^22^

All genetic variants used as instrumental variables (IV) were evaluated against the three core assumptions of MR: relevance, independence, and exclusion-restriction. Both individual-level and summary-level MR analyses were conducted accordingly, assuming that IVs affect the outcome solely through the exposure of interest and are independent of confounders (**Supplemental Figure S1A**).^13^ To assess instrument strength and minimize weak instrument bias, we calculated F-statistics, conditional F-statistics for multivariable models, and the proportion of variance explained (R²). SNP information, including identifiers, genomic positions, effect alleles and frequencies, and association estimates for each exposure, is provided in **Supplemental Tables S3–S5**.

### Mendelian randomization analyses and bidirectional causal assessment

Associations between the calculated GRS for ECG-aging and AF risk were examined using logistic regression as a case–control analysis, adjusted for genotype batch, assessment center, and 10 genetic principal components, in addition to the covariables used in the observational analyses. To illustrate risk stratification, AF incidence was compared between the top 10% and bottom 90% of the GRS distribution using the Kaplan–Meier method.

To strengthen and replicate causal inference, two-sample MR analyses were conducted using summary-level data from two large, ancestry-specific GWAS for AF, including the FinnGen study, which is independent of the UK Biobank-based exposure dataset.^23,24^ This approach increased statistical power by integrating independent genetic associations for exposure and outcome.^25^ All instrumental SNPs were aligned to GRCh37 genomic coordinates, with harmonization of strand orientation and effect alleles. Palindromic SNPs and those missing in the outcome GWAS were excluded, and no proxies were used. Primary MR estimates were obtained using the inverse-variance weighted (IVW) method with multiplicative random effects.^26^ To account for potential pleiotropy, complementary MR methods included MR-Egger, weighted median, MR-RAPS (Robust Adjusted Profile Score), and MR-PRESSO (Mendelian Randomization Pleiotropy RESidual Sum and Outlier). Details of complementary MR methods are provided in the **Supplemental Methods**.

To assess the possibility of reverse causality between ECG-aging and AF, bidirectional MR was performed treating AF as the exposure and ECG-aging as the outcome (**Supplemental Figure S1B**). This approach aimed to assess whether a genetic predisposition to AF might lead to accelerated ECG-aging. IVW served as the primary method, with sensitivity analyses consistent with the main MR approach. Steiger filtering was applied to enhance the reliability of causal direction inference by excluding SNPs more strongly associated with the outcome than the exposure (**Supplemental Methods**).

### Multivariable, mediation, and non-linear Mendelian randomization analyses

To investigate potential causal pathways linking ECG-aging to AF, we performed multivariable MR (MVMR) and two-step MR-based mediation analyses using summary-level data (**Supplemental Figure S1C**).^27^ Candidate mediators known to be associated with AF were first assessed via univariable MR to determine their association with genetically predicted ECG-aging. The total effect of ECG-aging on AF was estimated using univariable MR, and mediators with significant associations were included in MVMR to estimate the independent direct effects of ECG-aging and the mediator on AF, using distinct genetic instruments to minimize pleiotropy. The indirect effect was derived as the product of the effect of ECG-aging on the mediator and the mediator’s effect on AF conditional on ECG-aging, with standard errors calculated using the delta method. The proportion mediated was calculated as the ratio of indirect to total effect. All MR estimates represented the average causal effect per 1-SD increase in the exposure. All summary-level MR analyses were conducted using the “MendelianRandomization” and “TwoSampleMR” R packages.^28,29^

We performed non-linear MR leveraging individual-level UK Biobank data to examine potential non-uniform associations or differential risks across levels of the exposure between genetically predicted ECG-aging and AF risk. Using the “nlmr” R package, we applied both fractional polynomial and piecewise linear methods to estimate localized average causal effects across deciles of the residual (instrument-free) exposure.^30^ Further methodological details are provided in the **Supplemental Methods**.

### Complementary and sensitivity analyses

To extend the primary AF-focused analyses, complementary analyses examined associations between ECG-aging and CMR-derived structural and functional parameters using observational and MR approaches. Sensitivity two-sample MR analyses using AF GWAS summary data excluding UK Biobank participants were performed to address potential sample overlap and enhance robustness of main MR findings. Additional details of complementary and sensitivity analyses are provided in the **Supplemental Methods**.

## Results

### Clinical characteristics and cardiac imaging measurements

Baseline characteristics of the 35,090 participants are summarized in **Table 1**. Individuals in higher AI-ECG age gap quartiles were chronologically younger but exhibited progressively larger positive age gaps. Compared with lower quartiles, they had modestly lower socioeconomic deprivation, higher BMI, slightly lower systolic but higher diastolic BP, greater prevalence of current smoking and alcohol intake, and progressively reduced PA levels. Higher quartiles were also associated with higher eGFR and lower prevalence of dyslipidemia. Interestingly, despite a greater burden of heart failure (HF) and some other cardiovascular comorbidities, conventional AF risk scores (CHARGE-AF and HARMS2-AF) declined across quartiles, suggesting that the AI-ECG age gap provides incremental risk information beyond established clinical risk models.

**Table 1.** Clinical characteristics of the study cohort.

|  | AI-ECG age gap quartiles |  |  |  |  | P-value* |
| --- | --- | --- | --- | --- | --- | --- |
|  | Total<br>(N = 35090) | Quartile 1<br>(N = 8773) | Quartile 2<br>(N = 8773) | Quartile 3<br>(N = 8772) | Quartile 4<br>(N = 8772) |  |
| **Chronological age (years)** | 64.0 ± 7.8 | 68.4 ± 6.3 | 65.2 ± 7.6 | 62.8 ± 7.7 | 59.4 ± 6.5 | <0.001 |
| **AI-ECG age (years)** | 49.3 ± 6.4 | 48.5 ± 6.1 | 49.2 ± 6.3 | 49.6 ± 6.5 | 50.1 ± 6.6 | <0.001 |
| **AI-ECG age gap (years)** | -0.0 ± 6.2 | -7.9 ± 3.2 | -2.0 ± 1.2 | 1.9 ± 1.2 | 7.8 ± 3.1 | <0.001 |
| **Sex** |  |  |  |  |  | 0.87 |
| Female | 18162 (51.8%) | 4512 (51.4%) | 4533 (51.7%) | 4561 (52.0%) | 4556 (51.9%) |  |
| Male | 16928 (48.2%) | 4261 (48.6%) | 4240 (48.3%) | 4211 (48.0%) | 4216 (48.1%) |  |
| **Race** |  |  |  |  |  | 1.00 |
| White | 35090 (100.0%) | 8773 (100.0%) | 8773 (100.0%) | 8772 (100.0%) | 8772 (100.0%) |  |
| **Townsend Deprivation Index** | -2.0 ± 2.7 | -2.2 ± 2.5 | -2.0 ± 2.6 | -2.0 ± 2.7 | -1.8 ± 2.7 | <0.001 |
| **BMI (kg/m<sup>2</sup>)** | 26.6 ± 4.2 | 26.3 ± 4.0 | 26.3 ± 4.1 | 26.7 ± 4.3 | 27.0 ± 4.5 | <0.001 |
| Underweight | 153 (0.4%) | 45 (0.5%) | 36 (0.4%) | 43 (0.5%) | 29 (0.3%) |  |
| Normal | 13569 (39.1%) | 3481 (40.0%) | 3566 (41.0%) | 3327 (38.3%) | 3195 (37.0%) |  |
| Overweight | 15084 (43.4%) | 3844 (44.1%) | 3746 (43.1%) | 3781 (43.5%) | 3713 (43.0%) |  |
| Obesity | 5918 (17.0%) | 1339 (15.4%) | 1341 (15.4%) | 1532 (17.6%) | 1706 (19.7%) |  |
| **Systolic BP (mmHg)** | 135.7 ± 17.8 | 136.4 ± 17.9 | 135.9 ± 18.1 | 135.6 ± 17.9 | 134.9 ± 17.4 | <0.001 |
| **Diastolic BP (mmHg)** | 81.5 ± 9.9 | 80.5 ± 9.7 | 81.2 ± 9.8 | 81.8 ± 9.9 | 82.7 ± 10.1 | <0.001 |
| **Smoking status** |  |  |  |  |  | <0.001 |
| Never smoked | 21352 (61.0%) | 5295 (60.5%) | 5314 (60.7%) | 5319 (60.8%) | 5424 (62.0%) |  |
| Ex-smoking | 11550 (33.0%) | 3089 (35.3%) | 2975 (34.0%) | 2854 (32.6%) | 2632 (30.1%) |  |
| Current smoking | 2113 (6.0%) | 371 (4.2%) | 465 (5.3%) | 578 (6.6%) | 699 (8.0%) |  |
| **Drinking status** |  |  |  |  |  | 0.013 |
| Lifetime abstinence | 717 (2.0%) | 215 (2.5%) | 192 (2.2%) | 146 (1.7%) | 164 (1.9%) |  |
| Former drinking | 719 (2.0%) | 176 (2.0%) | 187 (2.1%) | 178 (2.0%) | 178 (2.0%) |  |
| Currently drinking | 33647 (95.9%) | 8381 (95.5%) | 8392 (95.7%) | 8447 (96.3%) | 8427 (96.1%) |  |
| **Physical activity (MET-min/week)** | 2484.1 ± 2433.6 | 2589.6 ± 2445.0 | 2561.3 ± 2513.2 | 2469.4 ± 2478.4 | 2317.9 ± 2284.3 | <0.001 |
| **Comorbidities** |  |  |  |  |  |  |
| Hypertension | 7204 (20.5%) | 1847 (21.1%) | 1797 (20.5%) | 1807 (20.6%) | 1753 (20.0%) | 0.38 |
| Diabetes mellitus | 873 (2.5%) | 235 (2.7%) | 221 (2.5%) | 205 (2.3%) | 212 (2.4%) | 0.50 |
| Dyslipidemia | 3606 (10.3%) | 1126 (12.8%) | 942 (10.7%) | 876 (10.0%) | 662 (7.5%) | <0.001 |
| Myocardial infarction | 428 (1.2%) | 122 (1.4%) | 113 (1.3%) | 114 (1.3%) | 79 (0.9%) | 0.016 |
| Heart failure | 727 (2.1%) | 138 (1.6%) | 188 (2.1%) | 177 (2.0%) | 224 (2.6%) | <0.001 |
| Peripheral arterial disease | 18 (0.1%) | 5 (0.1%) | 4 (0.0%) | 6 (0.1%) | 3 (0.0%) | 0.77 |
| Ischemic stroke | 162 (0.5%) | 46 (0.5%) | 43 (0.5%) | 41 (0.5%) | 32 (0.4%) | 0.44 |
| Venous thromboembolism | 221 (0.6%) | 59 (0.7%) | 60 (0.7%) | 50 (0.6%) | 52 (0.6%) | 0.72 |
| Chronic kidney disease | 189 (0.5%) | 59 (0.7%) | 56 (0.6%) | 43 (0.5%) | 31 (0.4%) | 0.014 |
| Sleep apnea | 151 (0.4%) | 31 (0.4%) | 35 (0.4%) | 44 (0.5%) | 41 (0.5%) | 0.43 |
| **Laboratory finding** |  |  |  |  |  |  |
| Hemoglobin A1c (%) | 5.4 ± 0.5 | 5.4 ± 0.5 | 5.4 ± 0.5 | 5.3 ± 0.5 | 5.3 ± 0.5 | <0.001 |
| Total cholesterol (mmol/L) | 5.7 ± 1.1 | 5.8 ± 1.1 | 5.8 ± 1.1 | 5.7 ± 1.1 | 5.7 ± 1.0 | <0.001 |
| LDL cholesterol (mmol/L) | 3.6 ± 0.8 | 3.6 ± 0.9 | 3.6 ± 0.8 | 3.6 ± 0.8 | 3.5 ± 0.8 | <0.001 |
| eGFR (mL/min/1.73m<sup>2</sup>) | 91.3 ± 12.1 | 88.2 ± 11.6 | 90.5 ± 12.0 | 92.1 ± 11.9 | 94.5 ± 12.1 | <0.001 |
| **CHARGE-AF score<sup>†</sup>** | 11.9 ± 0.9 | 12.4 ± 0.8 | 12.0 ± 1.0 | 11.8 ± 1.0 | 11.5 ± 0.8 | <0.001 |
| **HARMS2-AF score<sup>‡</sup>** | 4.0 ± 2.6 | 4.4 ± 2.5 | 4.1 ± 2.7 | 3.9 ± 2.7 | 3.5 ± 2.6 | <0.001 |
| **Follow-up time (years)** | 3.0 ± 1.6 | 2.9 ± 1.5 | 3.0 ± 1.5 | 3.0 ± 1.5 | 3.1 ± 1.6 | <0.001 |

Continuous variables are presented as mean ± SD and categorical variables are presented as number (percentage).
\* P-values for comparing the quartile groups were calculated using Pearson's chi-squared test or Fisher's exact test for categorical variables, and one-way analysis of variance (ANOVA) or the Kruskal–Wallis test for continuous variables, as appropriate.
<sup>†</sup> CHARGE-AF risk score was calculated using the following formula: $0.508 \times \text{age (per 5 years)} + 0.248 \times \text{height (per 10 cm)} + 0.115 \times \text{weight (per 15 kg)} + 0.197 \times \text{systolic BP (per 20 mmHg)} - 0.101 \times \text{diastolic BP (per 10 mmHg)} + 0.359 \times \text{current smoker} + 0.349 \times \text{hypertension} + 0.237 \times \text{diabetes} + 0.701 \times \text{congestive heart failure} + 0.496 \times \text{myocardial infarction}$ .
<sup>‡</sup> HARMS2-AF risk score was calculated using the following formula: $4 \times \text{hypertension} + 1 \times \text{age 60–64 years} + 2 \times \text{age} \geq 65 \text{ years} + 1 \times \text{BMI} \geq 30 \text{ kg/m}^2 + 2 \times \text{male sex} + 2 \times \text{sleep apnea} + 1 \times \text{alcohol intake of 7–14 standard drinks/week} + 2 \times \text{alcohol intake} \geq 15 \text{ drinks/week} + 1 \times \text{current smoker}$ .
Abbreviations: AF, atrial fibrillation; AI, artificial intelligence; BMI, body mass index; BP, blood pressure; ECG, electrocardiogram; eGFR, estimated glomerular filtration rate; LDL, low-density lipoprotein; MET, metabolic equivalent task; SD, standard deviation.

In comparisons of CMR parameters across AI-ECG age gap quartiles (**Supplemental Table S6**), higher quartiles were associated with progressively larger left atrial (LA) and left ventricular (LV) chamber volumes, increased myocardial mass, and greater pericardial adipose tissue (PAT) area. Indexed LA and LV volumes showed consistent upward trends across quartiles. Higher AI-ECG age gaps were also linked to subtle LV dysfunction, including mildly reduced ejection fraction (EF) and attenuated strain parameters.

### Observational associations between electrocardiographic aging and AF

During a mean follow-up of 3.01 years, 488 participants (1.39%) developed incident AF. Adjusted event rates increased progressively from 3.93 to 10.70 per 1,000 PY across AI-ECG age gap quartiles. In multivariable Cox regression, the highest quartile showed more than a twofold higher risk of incident AF compared with the lowest (HR 2.38 [95% CI 1.80–3.14]). Full results are provided in **Supplemental Table S7**. Adjusted cumulative incidence curves demonstrated consistent stepwise increases in AF incidence with higher quartiles (**Supplemental Figure S2**). These associations remained robust and consistent after accounting for the competing risk of death (**Supplemental Table S8**). Interestingly, participants with prevalent AF—excluded from the main analysis—displayed significantly higher AI-ECG age gaps compared with those without AF history (**Supplemental Figure S3**).

### Causal and directional associations between electrocardiographic aging and AF

Genetic instruments for the AI-ECG age gap demonstrated strong relevance (F-statistic of 41.7) and explained 21.9% of the phenotypic variance. Concordant with observational associations of higher AI-ECG age gaps with increased AF risk (HR 1.43 [95% CI, 1.29–1.59] per 1-SD increase), GRS-based MR analyses additionally provided genetic evidence supporting a causal link (OR 1.06 [95% CI, 1.01–1.12]; **Figure 2**). Stratified analysis further showed that individuals in the top 10% of the GRS distribution had a substantially higher cumulative incidence of AF than the lower 90% (**Supplemental Figure S4**). Two-sample MR analyses using two large, independent GWAS datasets further reinforced this causal relationship. Genetically predicted ECG-aging was associated with increased AF risk in both Nielsen et al. (Odds ratio [OR] 1.13 [95% CI, 1.02–1.25]) and the FinnGen study (OR 1.13 [95% CI, 1.02–1.25]). Complementary MR analyses yielded directionally consistent estimates across methods, with no evidence of horizontal pleiotropy and no indication of influential outlier–driven heterogeneity, supporting the robustness of the findings (**Figure 2 and Supplemental Table S9**).

**Figure 2.**
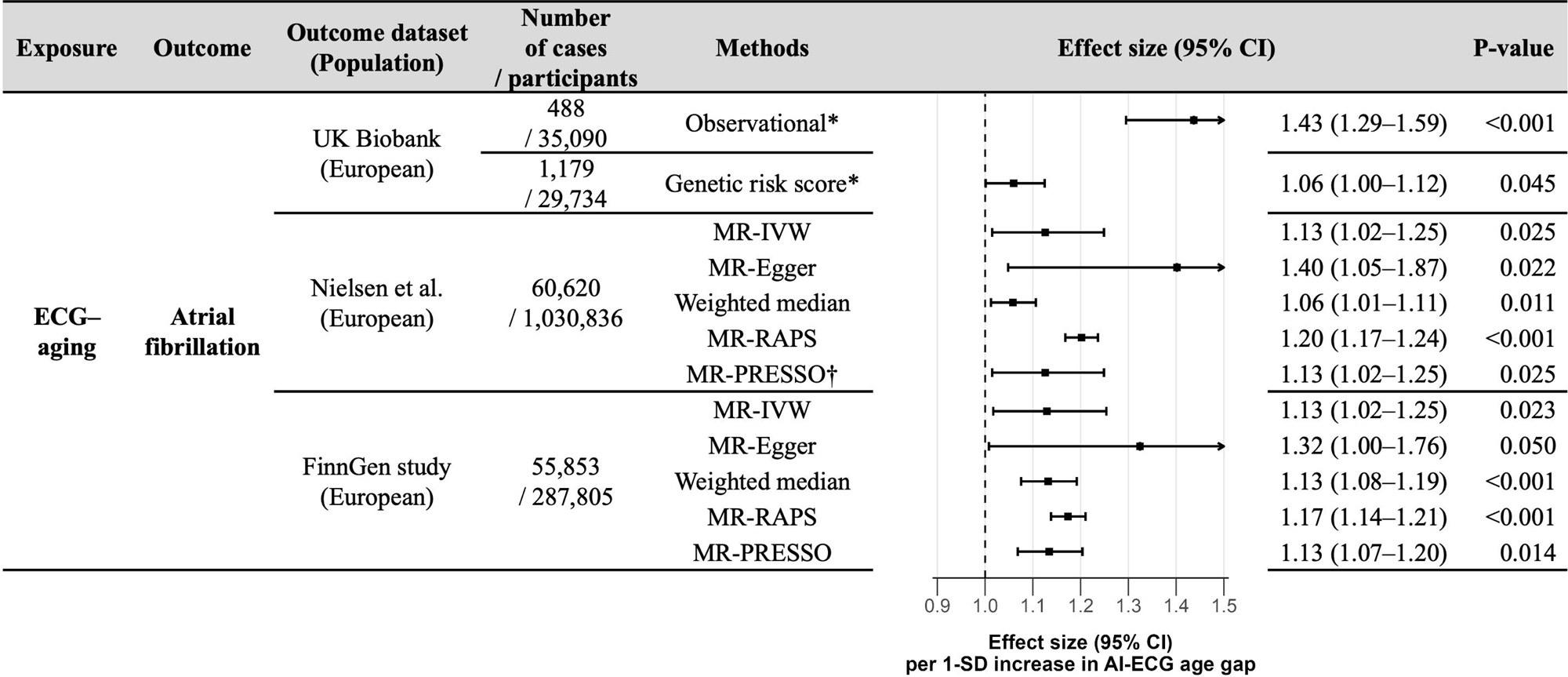
Observational and Mendelian randomization analyses for the association between ECG-aging and AF risk across multiple independent datasets Effect estimates of measured and genetically predicted ECG-aging on AF risk were derived from multivariable Cox regression, individual-level GRS-based MR, and summary-level two-sample MR. * Effect estimates from both the observational and genetic analyses were adjusted for covariables as detailed in the Methods section. † The MR-PRESSO global test (Global p-value = 0.23) did not detect any significant outliers in the genetic instrument, and no correction was necessary. As a result, the causal estimate calculated by MR-PRESSO was the same as that by the IVW method. Abbreviations: AF, atrial fibrillation; AI, artificial intelligence; CI, confidence interval; ECG, electrocardiogram; ECG-aging, electrocardiographic aging; IVW, inverse-variance weighted; MR, Mendelian randomization; PRESSO, pleiotropy residual sum and outlier; GRS, genetic risk score; RAPS, robust adjusted profile score; SD, standard deviation.

In bidirectional analyses (**Figure 3**), genetically predicted AF was initially associated with accelerated ECG-aging (OR 1.37 [95% CI, 1.11–1.71]), with sensitivity MR estimates—though not significant for MR-Egger—remaining directionally consistent, suggesting a possible reverse causal effect. After Steiger filtering, however, these reverse associations were attenuated and no longer statistically significant across MR methods (**Supplemental Table S10**). By contrast, the forward direction remained robust, reinforcing ECG-aging to AF as the primary causal pathway.

**Figure 3.**
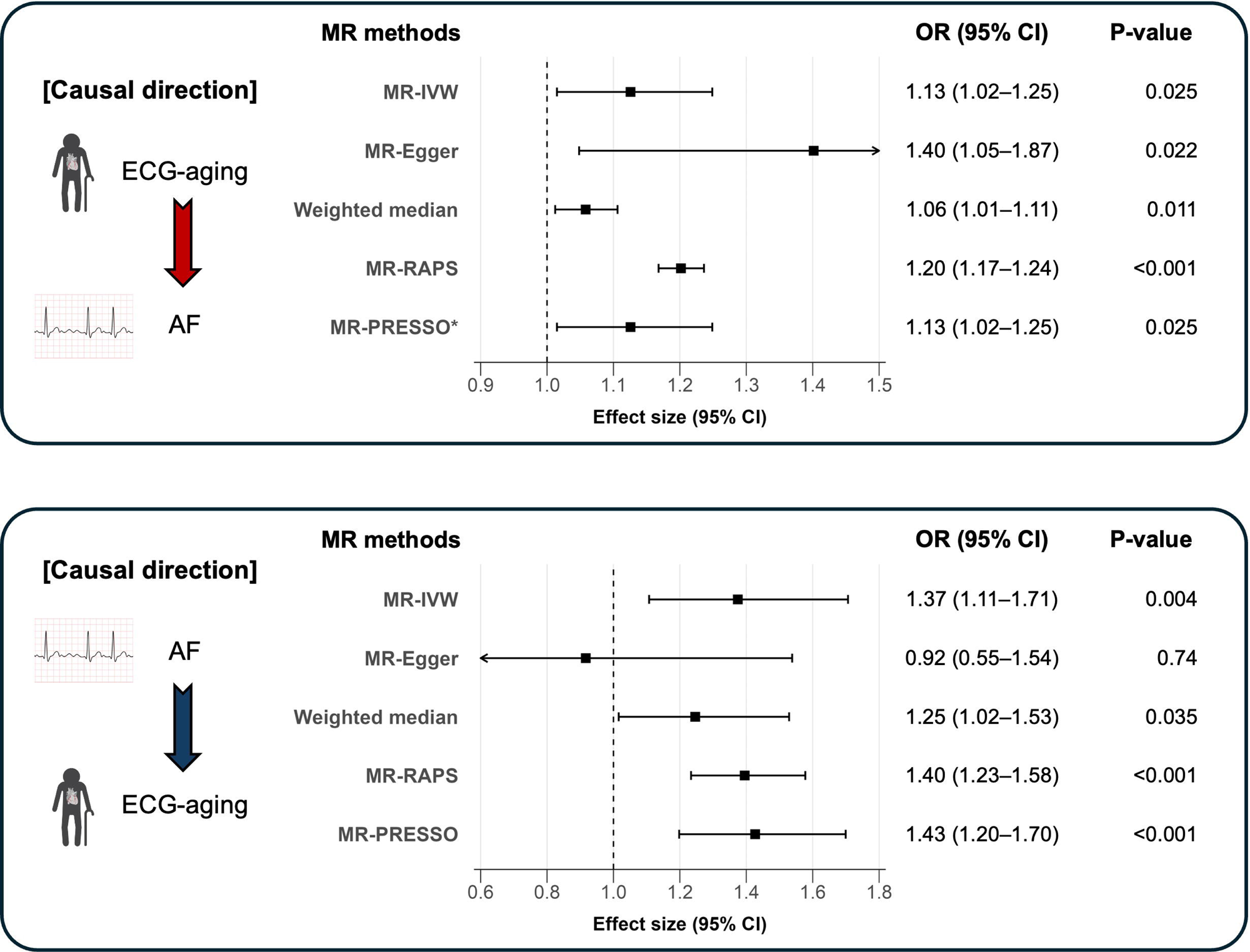
Bidirectional Mendelian randomization analyses of the causal relationship between ECG-aging and AF risk Bi-directional summary-level MR results showing causal estimates from a 1-SD increase in the AI-ECG age gap on AF risk, and from a 1-unit increase in the log-odds of AF on the AI-ECG age gap. Various MR methods were used for sensitivity analyses. The unit of the summary statistics for AF was aligned with the relative risk scale reported in the previous GWAS meta-analysis. GWAS summary data for ECG-aging and AF (Nielsen et al.) were alternately used as exposure and outcome in the bidirectional framework. * The MR-PRESSO global test (Global p-value = 0.23) did not detect any significant outliers in the genetic instrument, and no correction was necessary. As a result, the causal estimate calculated by MR-PRESSO was the same as that by the IVW method. Abbreviations: AF, atrial fibrillation; AI, artificial intelligence; CI, confidence interval; ECG, electrocardiogram; ECG-aging, electrocardiographic aging; GWAS, genome-wide association study; IVW, inverse-variance weighted; MR, Mendelian randomization; PRESSO, pleiotropy residual sum and outlier; RAPS, robust adjusted profile score; SD, standard deviation.

### Multivariable, mediation, and non-linear Mendelian randomization results

In univariable MR analyses, among candidate mediators known to be associated with AF, only HF showed a significant association with genetically predicted ECG-aging (OR 1.06 [95% CI, 1.01–1.12]), whereas no significant causal associations were observed for the other candidate mediators (**Table 2**). HF further exerted a substantial causal effect on AF risk (OR 3.85 [95% CI, 1.45–10.22]). In multivariable MR simultaneously modeling ECG-aging and HF (conditional F-statistics of 19.1 and 42.6, respectively) to examine mediation via HF, both exposures retained significant and independent associations with AF (ECG-aging: OR 1.07 [95% CI, 1.02–1.12]; HF: OR 3.79 [95% CI, 1.43–10.04]). Mediation analysis further demonstrated that the indirect effect of ECG-aging on AF through HF was significant (OR 1.09 [95% CI, 1.01–1.20]), accounting for approximately 70% of the total effect. These findings highlight HF as a key intermediary pathway through which accelerated ECG-aging contributes to AF risk, while also supporting a direct effect independent of HF.

**Table 2.**
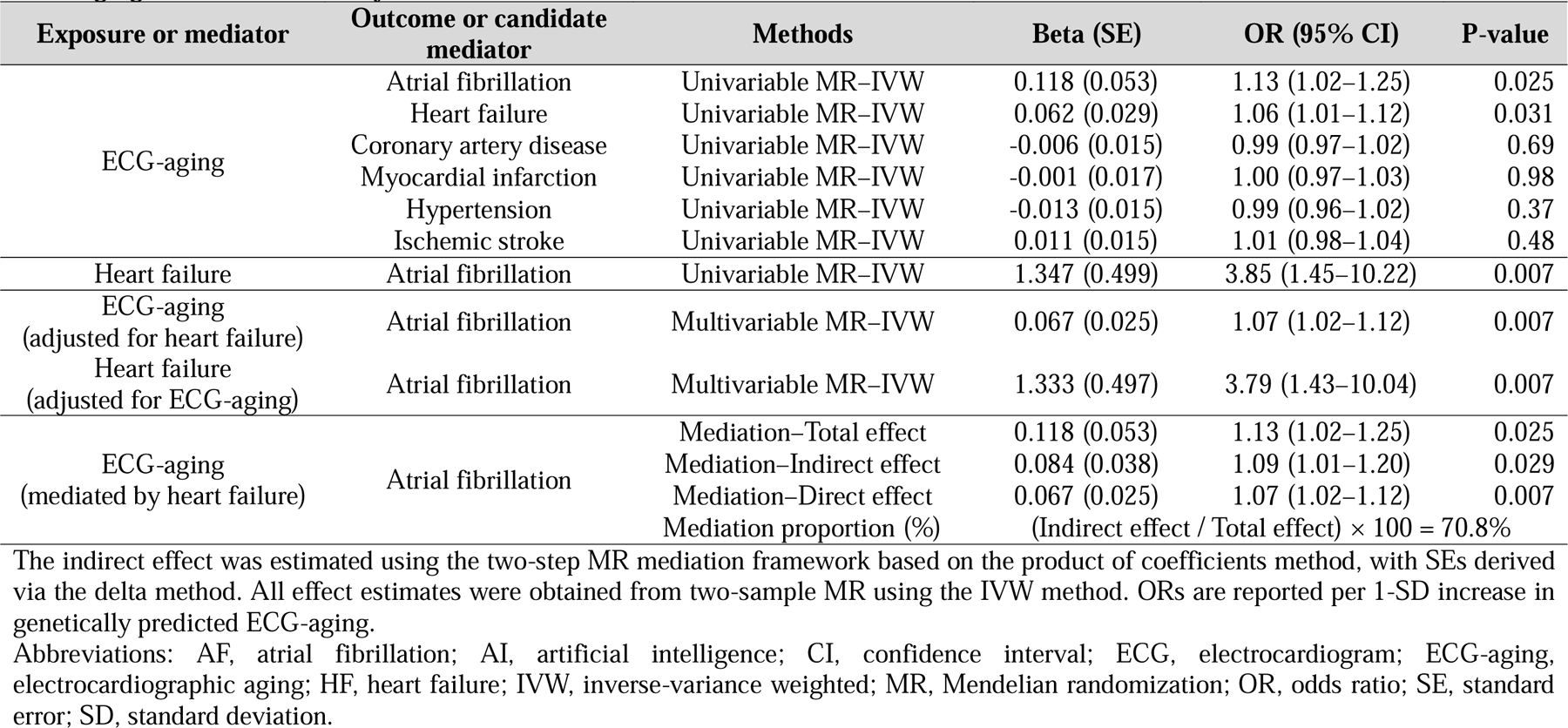
Multivariable Mendelian randomization and mediation analyses of the causal association between genetically predicted ECG-aging and AF, mediated by HF.

| Exposure or mediator | Outcome or candidate mediator | Methods | Beta (SE) | OR (95% CI) | P-value |
| --- | --- | --- | --- | --- | --- |
| ECG-aging | Atrial fibrillation | Univariable MR–IVW | 0.118 (0.053) | 1.13 (1.02–1.25) | 0.025 |
|  | Heart failure | Univariable MR–IVW | 0.062 (0.029) | 1.06 (1.01–1.12) | 0.031 |
|  | Coronary artery disease | Univariable MR–IVW | -0.006 (0.015) | 0.99 (0.97–1.02) | 0.69 |
|  | Myocardial infarction | Univariable MR–IVW | -0.001 (0.017) | 1.00 (0.97–1.03) | 0.98 |
|  | Hypertension | Univariable MR–IVW | -0.013 (0.015) | 0.99 (0.96–1.02) | 0.37 |
|  | Ischemic stroke | Univariable MR–IVW | 0.011 (0.015) | 1.01 (0.98–1.04) | 0.48 |
| Heart failure | Atrial fibrillation | Univariable MR–IVW | 1.347 (0.499) | 3.85 (1.45–10.22) | 0.007 |
| ECG-aging<br>(adjusted for heart failure) | Atrial fibrillation | Multivariable MR–IVW | 0.067 (0.025) | 1.07 (1.02–1.12) | 0.007 |
| Heart failure<br>(adjusted for ECG-aging) | Atrial fibrillation | Multivariable MR–IVW | 1.333 (0.497) | 3.79 (1.43–10.04) | 0.007 |
| ECG-aging<br>(mediated by heart failure) | Atrial fibrillation | Mediation–Total effect | 0.118 (0.053) | 1.13 (1.02–1.25) | 0.025 |
|  |  | Mediation–Indirect effect | 0.084 (0.038) | 1.09 (1.01–1.20) | 0.029 |
|  |  | Mediation–Direct effect | 0.067 (0.025) | 1.07 (1.02–1.12) | 0.007 |
|  |  | Mediation proportion (%) | (Indirect effect / Total effect) × 100 = 70.8% |  |  |

The indirect effect was estimated using the two-step MR mediation framework based on the product of coefficients method, with SEs derived via the delta method. All effect estimates were obtained from two-sample MR using the IVW method. ORs are reported per 1-SD increase in genetically predicted ECG-aging.
Abbreviations: AF, atrial fibrillation; AI, artificial intelligence; CI, confidence interval; ECG, electrocardiogram; ECG-aging, electrocardiographic aging; HF, heart failure; IVW, inverse-variance weighted; MR, Mendelian randomization; OR, odds ratio; SE, standard error; SD, standard deviation.

Non-linear MR revealed a statistically significant effect of genetically predicted ECG-aging on AF risk (**Figure 4**). Fractional polynomial modeling showed a modest curvilinear trend, with a marginally significant quadratic term (p = 0.046), suggesting that the effect of genetically predicted ECG-aging on AF may intensify at higher levels of ECG-aging. The piecewise MR analysis yielded consistent findings, showing a gradual rise in AF risk beyond the reference range, although the overall test for non-linearity was not statistically significant.

**Figure 4.**
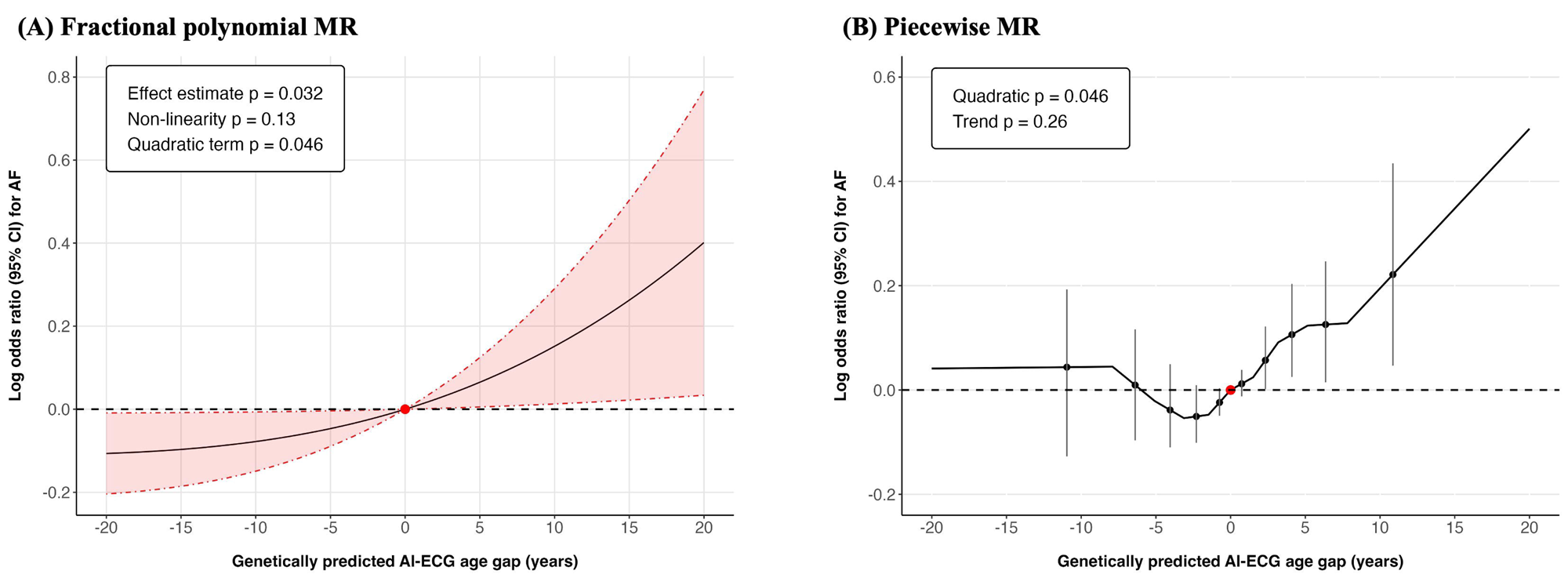
Nonlinear Mendelian randomization analyses on the relationship between genetically predicted ECG-aging and AF risk (A) Fractional polynomial MR analysis illustrating the estimated log odds for AF across the continuum of genetically predicted AI-ECG age gap. (B) Piecewise MR analysis displaying localized average causal effects across strata of the exposure distribution. The gradient at each value of genetically predicted AI-ECG age gap corresponds directly to the localized average causal effect within that residualized exposure stratum. In both panels, nonlinear associations were evaluated using quadratic and trend tests, with the relevant p-values shown in each figure. Solid lines denote point estimates, and shaded areas (A) or vertical error bars (B) represent 95% CIs. Abbreviations: AF, atrial fibrillation; AI, artificial intelligence; CI, confidence interval; ECG, electrocardiogram; MR, Mendelian randomization.

### Associations between electrocardiographic aging and cardiac structure and function

Across CMR-derived structural and functional measurements, higher AI-ECG age gaps were associated with adverse cardiac remodeling in observational analyses, including larger LA and LV volumes, reduced LA emptying, increased myocardial mass and wall thickness, and greater PAT area (**Supplemental Table S11**). GRS-based genetic associations revealed a more selective pattern (**Supplemental Table S12**). While most volumetric and functional measures showed no significant genetic effects, myocardial mass and LV wall thickness remained significantly associated, indicating LV hypertrophic remodeling among individuals genetically predisposed to accelerated ECG-aging. These concordant observational and genetic signals emphasize structural hypertrophy as the primary remodeling feature linked to ECG-aging (**Supplemental Figure S5**). Consistent with these findings, non-linear MR analyses focusing on LV hypertrophic remodeling–related traits demonstrated largely linear associations across the exposure distribution, with wall thickness showing modest curvature characterized by localized upward shifts at higher levels of the exposure (**Supplemental Figure S6–S7**).

### Sensitivity analyses

In sensitivity two-sample MR analyses using AF GWAS summary statistics excluding UK Biobank participants, genetically predicted ECG-aging remained consistently associated with increased AF risk across multiple MR methods, supporting the robustness of our main findings (**Supplemental Table S13**). Scatter plots further demonstrated consistent SNP-specific causal estimates in both causal directions, including analyses using AF GWAS datasets excluding UK Biobank participants (**Supplemental Figures S8–S9**). Leave-one-out sensitivity analyses showed that no single variant disproportionately influenced the causal estimates in either direction, confirming the stability and concordance of the findings across all outcome datasets (**Supplemental Figures S10–S11**).

## Discussion

### Main findings

In this study, we demonstrated a consistent and robust causal association between ECG-aging, as reflected by the AI-ECG age gap, and AF risk using both observational analyses and various MR frameworks. Observationally, higher AI-ECG age gaps were linked to an increased risk of incident AF, findings that were directionally concordant with genetic analyses. Genetically predicted ECG-aging showed a causal relationship with AF in individual-level GRS-based MR and was replicated in independent two-sample MR using large external GWAS summary datasets. Sensitivity analyses, including complementary MR approaches, bidirectional testing, and non-linear MR, supported the robustness and directionality of this relationship, while suggesting a possible subtle curvilinear association across the AI-ECG age gap spectrum. Notably, bidirectional MR testing initially suggested effects in both directions, but Steiger filtering confirmed ECG-aging to AF as the predominant causal pathway. MVMR and mediation analyses suggested that a substantial proportion of the ECG-aging–AF relationship may be mediated through HF. Beyond AF risk, both measured and genetically predicted ECG-aging were associated with adverse cardiac structural and functional remodeling on CMR data, suggesting that accelerated ECG-aging may reflect underlying myocardial alterations predisposing to atrial arrhythmogenesis.

Consequently, leveraging genetic instruments in comprehensive MR analyses, we mitigated limitations of observational approaches related to time-varying exposure measurement, thereby strengthening causal inference for its role in AF development.

### Clinical and therapeutic implications of accelerated electrocardiographic aging

This study provides the first genetic evidence supporting a potential causal relationship between AI-derived ECG-based aging parameters and the risk of AF. By integrating deep learning-based phenotypes from routine 12-lead ECGs with various MR frameworks leveraging robust genetic instruments, our findings extend prior observational work by demonstrating that accelerated ECG-aging is not merely a correlate of cardiovascular risk but a potential causal factor in AF development. These results support the hypothesis that electrophysiological alterations captured by AI reflect biologically meaningful deviations in cardiac aging that contribute to atrial remodeling and arrhythmogenesis beyond shared risk factors or subclinical disease.^31^

If ECG-aging plays a direct causal role in AF development, it may function not only as a prognostic biomarker but also as a potential interventional target. Unlike many emerging digital biomarkers that have faced limited clinical adoption due to the absence of clear biological rationale or causal evidence, our MR findings provide genetic support that accelerated ECG-aging lies on a causal pathway to AF. Similar to other modifiable causal risk factors such as obesity, hypertension, or diabetes, ECG-aging may represent an early, actionable state of electrophysiological vulnerability preceding overt AF.^32^ This causal anchoring supports the concept that identifying high-risk individuals through AI-ECG age screening could enable upstream AF prevention strategies, including intensive risk factor modification, early rhythm control, or novel interventions aimed at preserving atrial electrical integrity.

### Pathophysiological insights into AF from electrocardiographic aging

Our results support the concept that AF may represent a clinical manifestation of accelerated cardiac aging, particularly involving structural and electrophysiological vulnerability. Both observed and genetically predicted ECG-aging were associated with adverse cardiac remodeling on CMR data, suggesting a plausible mechanistic continuum from electrical aging to structural substrate predisposition for AF.^7^ In particular, increased LV mass and wall thickness—reflecting pressure overload, concentric hypertrophy, or interstitial fibrosis—can raise LV filling pressures and impose chronic stress on the LA, promoting LA remodeling and fibrosis that ultimately create a substrate for atrial arrhythmogenesis.^33,34^ Thus, larger AI-ECG age gaps likely capture not only electrophysiologic aging but also subclinical structural remodeling predisposing to AF.

Mediation MR analyses further implicated HF as a key intermediate phenotype, explaining approximately 70% of the ECG-aging–AF association. This pathway suggests that early electrophysiological decline drives subclinical myocardial dysfunction, ultimately increasing susceptibility to AF. The pronounced atrial-specific vulnerability to cardiac aging—likely mediated through tissue architecture, fibrosis, or ion-channel remodeling—aligns with established mechanisms in cardiovascular senescence.^35,36^ From a broader perspective, AF may be considered a cardiac manifestation of premature aging, particularly evident in individuals with accelerated electrophysiological decline. The LA, with its unique structural and molecular characteristics, may be particularly susceptible to age-related degeneration compared with other cardiac chambers.^35,37^ This chamber-specific vulnerability aligns with patterns observed in other organ systems, where certain tissues demonstrate greater sensitivity to age-related deterioration.

Collectively, these findings position ECG-aging as an upstream biomarker of atrial pathophysiology that bridges electrical, structural, and clinical facets of AF. As a genetically anchored risk signal, it refines risk stratification and unveils novel mechanistic pathways toward prevention, potentially enabling precision-targeted strategies aimed at mitigating electrophysiological aging beyond conventional risk factor management.

### Limitations

This study has several limitations. First, the median follow-up duration of approximately three years may be insufficient to fully assess long-term AF risk. Second, despite extensive complementary and sensitivity analyses aimed to minimize pleiotropy, the observed associations between ECG-aging and AF may still partly reflect shared genetic architectures and factors, or unmeasured pleiotropic pathways not fully accounted for by current MR methods, rather than a purely direct causal effect. Third, despite adequate F-statistics, the relatively small number of genome-wide significant SNPs raises the possibility of residual weak instrument bias that cannot be entirely ruled out. Fourth, partial sample overlap between datasets used for instrument derivation and outcome estimation in two-sample MR could bias causal effect estimates. To mitigate this concern, we conducted sensitivity analyses using GWAS summary data excluding UK Biobank participants and replicated MR findings in the independent FinnGen consortium, reducing the likelihood of false positives. Fifth, our analyses were restricted to individuals of European ancestry to minimize potential confounding by population stratification, which limits the applicability of our findings to other ancestries. Future studies in diverse ethnic populations will be essential to assess the generalizability of our findings. Finally, although individual-level GRS-based MR was limited to a single cohort, we strengthened the robustness of our findings by performing two-sample MR analyses using independent external GWAS datasets for AF. Further large-scale GWAS results beyond UK Biobank will be needed to validate and refine the genetic instrument for ECG-aging.

## Conclusions

This study provides robust and compelling genetic evidence supporting a causal association between ECG-aging, as captured by the AI-ECG age gap, and the risk of AF. Using multiple MR frameworks, we consistently demonstrated the validity and reliability of this causal inference across complementary analytic approaches. Our findings extend beyond prior observational studies by establishing ECG-aging as not only a prognostic marker but also a causally relevant, clinically actionable biomarker in AF pathogenesis and risk stratification. Integrating ECG-aging into AF risk prediction or screening may enable earlier identification of high-risk individuals for upstream preventive strategies.

## Acknowledgements

We gratefully acknowledge the participants and investigators of UK Biobank and the FinnGen consortium for providing invaluable data resources.

## Author contributions

The authors are entirely responsible for the study’s design and implementation, all analyses performed, the paper’s drafting and editing, and its final content.

## Data availability

The anonymized data used in this study can be requested in whole or in part by any qualified investigator for the purposes of replicating the analyses and results and will be made available pending ethics clearance and approval by authors and institutions of each of the sites the data are requested from. The UK Biobank data used in this study were obtained through the UK Biobank Consortium in accordance with the relevant data use agreements (https://biobank.ndph.ox.ac.uk/ukb/). GWAS summary-level data were obtained from publicly available repositories, including the GWAS Catalogue (https://www.ebi.ac.uk/gwas/), the IEU OpenGWAS project (https://gwas.mrcieu.ac.uk/), and FinnGen (https://www.finngen.fi/en).

