## Supplemental Materials for "Accelerated electrocardiographic aging of the heart as a risk determinant for atrial fibrillation: A Mendelian randomization study"

**Co-corresponding authors:**

**– Table of Contents –**

**Supplemental Methods**

**Supplemental Table S1.** Definitions and ICD-10 codes for diagnoses of comorbidities and study outcomes

**Supplemental Table S2.** Summary of GWAS data sources for exposure, mediator and outcome traits used in this study

**Supplemental Table S3.** List of genetic instrumental variables for ECG-aging in the Mendelian randomization analysis

**Supplemental Table S4.** List of genetic instrumental variables for AF in the Mendelian randomization analysis

**Supplemental Table S5.** List of genetic instrumental variables for HF in the Mendelian randomization analysis

**Supplemental Table S6.** Comparisons of CMR imaging measurements by AI-ECG age gap quartiles

**Supplemental Table S7.** Incidence and risk of AF across AI-ECG age gap quartiles

**Supplemental Table S8.** Risk of AF across AI-ECG age gap quartiles accounting for competing risk of death

**Supplemental Table S9.** Heterogeneity and horizontal pleiotropy assessments for Mendelian randomization analyses of ECG-aging and AF

**Supplemental Table S10.** Steiger-filtered bidirectional Mendelian randomization results of the causal association between ECG-aging and AF risk

**Supplemental Table S11.** Multivariable regression analyses of the associations between AI-ECG age gap and CMR imaging measurements

**Supplemental Table S12.** GRS-based Mendelian randomization analyses of associations between genetically predicted ECG-aging and CMR imaging measurements

**Supplemental Table S13.** Two-sample Mendelian randomization analyses of the causal association between ECG-aging and AF using UK Biobank–excluded outcome GWAS summary statistics

**Supplemental Figure S1.** Schematics of Mendelian randomization analyses performed in this study

**Supplemental Figure S2.** Adjusted cumulative incidence of AF according to AI-ECG age gap quartiles

**Supplemental Figure S3.** Differences in AI-ECG age gaps between participants with and without prevalent AF at baseline

**Supplemental Figure S4.** Cumulative incidence curves of AF stratified by GRS percentile groups

**Supplemental Figure S5.** Pyramid plot of observational and genetic associations between AI-ECG age gaps and CMR imaging measurements

**Supplemental Figure S6.** Fractional polynomial Mendelian randomization analysis of the association between genetically predicted ECG-aging and LV remodeling–related CMR traits

**Supplemental Figure S7.** Piecewise Mendelian randomization analysis of the association between genetically predicted ECG-aging and LV remodeling–related CMR traits

**Supplemental Figure S8.** Scatter plot comparing causal estimates of ECG-aging on AF across multiple Mendelian randomization methods

**Supplemental Figure S9.** Scatter plot comparing causal estimates of AF on ECG-aging across multiple Mendelian randomization methods

**Supplemental Figure S10.** Leave-one-out Mendelian randomization sensitivity analysis for the association between genetically predicted ECG-aging and AF risk

**Supplemental Figure S11.** Leave-one-out Mendelian randomization sensitivity analysis for the association between genetically predicted AF risk and ECG-aging

**Supplemental References**

**Supplemental Methods**

***Two-sample Mendelian randomization sensitivity analyses***

To evaluate the robustness of the primary inverse-variance weighted (IVW) estimates and assess the potential influence of horizontal pleiotropy, a series of complementary Mendelian randomization (MR) sensitivity analyses using well-established methodological frameworks were conducted. All analyses were performed using summary-level single-nucleotide polymorphism (SNP)-exposure and SNP-outcome associations harmonized across datasets. The MR-Egger method was used to detect and account for directional horizontal pleiotropy.^1^ This approach provides unbiased causal estimates even when all included variants exert pleiotropic effects, under the InSIDE (Instrument Strength Independent of Direct Effect) assumption.^2^ The MR-Egger intercept was evaluated as a formal test for directional pleiotropy. The weighted median estimator yields consistent causal estimates even if up to 50% of the weight in the analysis originates from invalid instruments, thereby offering robustness to violations of the exclusion-restriction assumption.^1^ MR-RAPS (Robust Adjusted Profile Score) was implemented to address potential bias from weak instruments and idiosyncratic pleiotropy. This method applies a robust likelihood-based framework suitable for settings in which instrument strength varies or when measurement error may induce regression dilution.^3^ The MR-PRESSO (Pleiotropy RESidual Sum and Outlier) method was applied to identify and correct for horizontal pleiotropic outliers using a global test and outlier-removal process.^4^ If significant outliers were detected, causal estimates were recalculated after outlier exclusion. Otherwise, the IVW estimates were retained as the MR-PRESSO-corrected estimates.

***Steiger filtering sensitivity analysis***

Steiger filtering was applied to evaluate the plausibility of the assumed causal direction and to reduce potential reverse causation bias in two-sample MR analyses.^5^ This method compares the variance explained by each genetic variant in the exposure and the outcome, excluding SNPs that explain more variance in the outcome than in the exposure and are therefore less likely to satisfy core MR assumptions that genetic instruments influence the outcome primarily through the exposure of interest.^5^ Implementation was performed using the steiger_filtering() function in the “TwoSampleMR” R package, which conducts SNP-level directionality tests based on summarized association statistics. SNPs failing the directionality criterion were excluded prior to rerunning MR analyses. This approach improves the reliability of causal inference by limiting bias from variants with stronger outcome associations, particularly in settings where bidirectional relationships or phenotype correlation may be present.

***Nonlinear Mendelian randomization analyses***

Non-linear MR analyses were conducted using individual-level UK Biobank data to assess potential departures from linearity and to examine heterogeneity in the causal associations of genetically predicted AI-ECG age gap (Artificial Intelligence-predicted ElectroCardioGram-derived age gap) with atrial fibrillation (AF) risk and cardiac magnetic resonance (CMR)-derived structural and functional imaging parameters across the exposure distribution. These analyses were performed using the “nlmr” R package, following previously established methodological frameworks.^6,7^ This approach enabled formal testing of non-linearity and flexible modeling of exposure–outcome relationships, providing deeper insights into the potential heterogeneity across the spectrum of the AI-ECG age gap.^8^

To avoid collider bias arising from stratification on a genetically influenced exposure, analyses were conducted within strata of residualized (instrument-free) exposure.^6,9^ Specifically, residual AI-ECG age gap was calculated by subtracting genetically predicted AI-ECG age gap from the observed AI-ECG age gap. This residualized exposure represents the expected AI-ECG age gap if all participants shared the same genotype and is therefore independent of the genetic instrument, allowing stratification without introducing overadjustment or collider bias.^7^ Participants were stratified into deciles of the residual AI-ECG age gap to maximize statistical power within strata. Within each decile, associations between the genetic risk score (GRS) and each outcome were estimated using logistic regression for binary outcomes and linear regression for continuous traits, adjusting for age, sex, genotype batch, assessment center, and 10 genetic principal components. Decile-specific localized average causal effects (LACE) were calculated as the ratio of the genetic association with the outcome to the genetic association with the exposure.^6,7^

Interval-specific LACE estimates were subsequently used to reconstruct the overall exposure–outcome relationship using both piecewise linear and fractional polynomial approaches. In the piecewise linear method, LACE estimates were treated as local slopes within each exposure stratum, allowing visualization of changes in causal effect magnitude across the exposure range.^9^ In fractional polynomial modeling, LACE estimates were meta-regressed against the mean exposure level in each stratum using predefined polynomial powers to flexibly model potential non-linear patterns.^6,9^ Model fit was formally compared with a linear model to test for evidence of non-linearity. Non-linearity was assessed using complementary statistical tests, including fractional polynomial non-linearity tests, quadratic trend tests, and Cochran’s Q tests for heterogeneity across exposure strata.^6^ Given that all estimates were derived within residualized exposure strata, interpretation focused on relative differences in causal slopes across the exposure distribution rather than absolute risk estimates.^7^

***Complementary and sensitivity analyses***

Complementary analyses were conducted to comprehensively evaluate the potential impact of measured and genetically predicted electrocardiographic aging (ECG-aging) on cardiac structural remodeling and functional changes using CMR-derived measurements, beyond AF risk. Observational associations were examined using multivariable regression models adjusted for relevant demographic and clinical covariables. GRS-based MR analyses were performed for CMR imaging traits using individual-level genotype and phenotype data, with causal estimates derived from instrumental variable regression frameworks. Integrated visualization of observational and genetic associations between AI-ECG age gaps and CMR imaging parameters was provided using pyramid plots to facilitate comparison of effect direction and magnitude across analytical frameworks. Non-linear MR analyses were additionally conducted to explore potential heterogeneity and non-linear exposure–outcome relationships using both fractional polynomial and piecewise approaches, as described in detail in the preceding section.

To enhance the robustness of our main summary-level MR findings for AF and to mitigate potential bias from sample overlap, sensitivity two-sample MR analyses were conducted using AF genome-wide association study (GWAS) summary statistics excluding UK Biobank participants, ensuring complete independence from the primary exposure dataset. These analyses followed the same instrumental variable and harmonization protocols as the primary framework and incorporated multiple MR methods, with complementary scatter plots of SNP-specific causal estimates and leave-one-out analyses to assess the influence of individual variants in both causal directions, including analyses based on an independent outcome dataset.

Additionally, as a sensitivity analysis, differences in AI-ECG age gaps between participants with and without prevalent AF at baseline were evaluated to contextualize observational findings.

**Supplemental Table S1. Definitions and ICD-10 codes for diagnoses of comorbidities and study outcomes**

| **Comorbidities** | **Definitions** | **ICD-10 codes**  **or related conditions** |
| --- | --- | --- |
| Atrial fibrillation | Defined using UK Biobank self-reported data or diagnostic codes* | Self-reported non-cancer illness code: 1471, 1483  ICD-10: I48 |
| Hypertension | Defined using UK Biobank self-reported data or diagnostic codes* | Self-reported non-cancer illness code: 1065, 1072  ICD-10: I10, I11, I12, I13, I15 |
| Diabetes mellitus | Defined using UK Biobank self-reported data or diagnostic codes* | Self-reported non-cancer illness code: 1220, 1222, 1223, 1521  ICD-10: E10, E11, E12, E13, E14 |
| Dyslipidemia | Defined using UK Biobank self-reported data or diagnostic codes* | Self-reported non-cancer illness code: 1473  ICD-10: E78 |
| Myocardial infarction | Defined using UK Biobank self-reported data or diagnostic codes* | Self-reported non-cancer illness code: 1075  ICD-10: I21, I22, I25.2 |
| Heart failure | Defined using UK Biobank self-reported data or diagnostic codes* | Self-reported non-cancer illness code: 1076  ICD-10: I11.0, I50, I97.1 |
| Peripheral arterial disease | Defined using UK Biobank self-reported data or diagnostic codes* | Self-reported non-cancer illness code: 1067, 1087  ICD-10: I70, I71 |
| Ischemic stroke | Defined using UK Biobank self-reported data or diagnostic codes* | Self-reported non-cancer illness code: 1583  ICD-10: I63, I64 |
| Venous thromboembolism | Defined using UK Biobank self-reported data or diagnostic codes* | Self-reported non-cancer illness code: 1068, 1093, 1094  ICD-10: I26, I80.1, I80.2, I80.3, I80.8, I80.9, I81, I82, I63.6, O22.3, O22.5, O87.1, O87.3 |
| Chronic kidney disease | Primarily defined using eGFR; otherwise, by self-reported illness codes or diagnostic codes* | eGFR <60 mL/min per 1.73 m2  Self-reported non-cancer illness code: 1192, 1194  ICD-10: N18, N19 |
| Sleep apnea | Defined using UK Biobank self-reported data or diagnostic codes* | Self-reported non-cancer illness code: 1123  ICD-10: G47.3 |
| **Study outcomes** | **Definitions** | **ICD-10 codes**  **or related conditions** |
| Atrial fibrillation | Defined using diagnostic codes* or related death without previous history of AF | ICD-10: I48 |

In the UK Biobank, the ICD-10 codes included converted 3-character ICD-10 codes from ICD-9-based diagnoses, using data-coding 1836.

* To improve diagnostic accuracy, diagnoses were defined as the presence of at least two outpatient records (equivalent to primary care records in the UK) or one or more hospital admissions corresponding to ICD-10 codes.

Abbreviations: AF, atrial fibrillation; eGFR, estimated glomerular filtration rate; HF, heart failure; ICD-10, international classification of diseases 10th revision.

**Supplemental Table S2. Summary of GWAS data sources for exposure, mediator and outcome traits used in this study**

| **Phenotypic traits** | **Type of traits** | **Definition of phenotype** | **Unit** | **Adjustments** | **Sample size (case) / Population** | **Study or consortium** | **PubMed ID* (publish year)** |
| --- | --- | --- | --- | --- | --- | --- | --- |
| ECG-aging | Exposure (or outcome in bi-directional MR framework) | AI-predicted ECG-age minus chronological age | SD increase in phenotype | Age, sex, the first ten genetic principal components, assessment center, and genotyping array | 31,475 /  European | UK Biobank | 37604819  (2023) |
| Atrial fibrillation | Outcome (or exposure in bi-directional MR framework) | Atrial fibrillation  (ICD-9/10 or 12-lead ECG) | Binary trait | Birth year or age, sex, and genetic principal components including study-specific covariates | 1,030,836  (60,620) /  European | The Nord-Trøndelag Health Study (HUNT) / deCODE / Michigan Genomics Initiative (MGI) / DiscovEHR / UK Biobank / AFGen Consortium | 30061737  (2018) |
| Atrial fibrillation / flutter (I9_AF) | Outcome | Atrial fibrillation or flutter (ICD-8/9/10 or KELA codes) | Binary trait | Age, sex, the first ten genetic principal components, and genotyping batch | 287,805  (55,853) /  European | FinnGen study (DF11) | 36653562  (2023) |
| Heart failure | Outcome or mediator | Heart failure of any etiology, regardless of LVEF (ICD-9/10 codes or clinical adjudication) | Binary trait | Age, sex, and genetic principal components including study-specific technical covariates | 977,323  (47,309) /  European | Atherosclerosis Risk in Communities (ARIC) / Cardiovascular Health Study (CHS) / UK Biobank / deCODE / BioBank Japan / BIOSTAT-CHF / and 20 additional cohorts | 31919418  (2020) |
| Coronary artery disease | Candidate mediator | Myocardial infarction, acute coronary syndrome, angina, or coronary revascularization (ICD-9/10, procedure codes, self-report, or clinical adjudication) | Binary trait | Age, sex, and genetic principal components including study-specific covariates (genotyping batch or recruitment center) | 1,165,690  (181,522) /  European | UK Biobank / deCODE genetics / The Nord-Trøndelag Health Study (HUNT) / TIMI Study / EPIC-CVD / Mass General Brigham Biobank / and 4 additional cohorts | 36474045  (2022) |
| Myocardial infarction | Candidate mediator | Myocardial infarction (ICD-10 codes, self-report, or physician-diagnosed) | Binary trait | Age, sex, genotyping array, and the first twenty genetic principal components | 639,221  (61,505) /  European | CARDIoGRAMplusC4D Consortium / UK Biobank | 33532862  (2021) |
| Hypertension (I9_HYPTENS) | Candidate mediator | Hypertension (ICD-8/9/10 or KELA codes) | Binary trait | Age, sex, the first ten genetic principal components, and genotyping batch | 453,657 (137,312) /  European | FinnGen study | 36653562  (2023) |
| Ischemic stroke | Candidate mediator | Ischemic stroke (WHO criteria, neuroimaging confirmation, clinical adjudication, or ICD-9/10 codes; with TOAST subtyping where available) | Binary trait | Age, sex, the first ten genetic principal components, genotyping array, and study-specific covariates | 440,328  (40,585) /  European | MEGASTROKE consortium | 29531354  (2018) |

* PubMed ID indicates PubMed identifier.

Abbreviations: AF, atrial fibrillation; AI, artificial intelligence; ECG, electrocardiogram; ECG-aging, electrocardiographic aging; GWAS, genome-wide association study; ICD, international classification of diseases; LVEF, left ventricular ejection fraction; MR, Mendelian randomization; SD, standard deviation; TOAST, trial of org 10172 in acute stroke treatment; WHO, world health organization.

**Supplemental Table S3. List of genetic instrumental variables for ECG-aging in the Mendelian randomization analysis**

| **SNP** | **Chromosome** | **Position* (GRCh37)** | **Gene** | **Effect allele** | **Other allele** | **Effect allele frequency** | **Beta** | **Standard error** | **P-value** |
| --- | --- | --- | --- | --- | --- | --- | --- | --- | --- |
| rs4671961 | 2 | 54571459 | EML6 | A | G | 0.32 | 0.222 | 0.039 | 9.1x10^-9^ |
| rs11902709 | 2 | 178743480 | TTN | T | C | 0.05 | 0.505 | 0.084 | 1.8x10^-9^ |
| rs7373065 | 3 | 38668824 | SCN5A | C | T | 0.02 | -0.926 | 0.137 | 1.2x10^-11^ |
| rs35430511 | 4 | 113465982 | CAMK2D | C | T | 0.25 | 0.274 | 0.041 | 2.5x10^-11^ |
| rs147790633 | 10 | 73687824 | MYOZ1 | C | T | 0.14 | -0.369 | 0.052 | 1.2x10^-12^ |
| rs60820984 | 10 | 73879820 | CAMK2G | T | C | 0.19 | -0.279 | 0.046 | 1.8x10^-9^ |
| rs7132327 | 12 | 114943266 | TBX3 | C | T | 0.27 | 0.251 | 0.040 | 5.4x10^-10^ |
| rs35866366 | 14 | 71382468 | SIPA1L1 | G | A | 0.24 | 0.288 | 0.041 | 3.4x10^-12^ |

* Genomic positions for all listed variants are reported based on the GRCh37/hg19 reference build.

Abbreviations: ECG-aging, electrocardiographic aging; GRCh37, Genome Reference Consortium human genome build 37; hg19, human genome version 19; SNP, single nucleotide polymorphism.

**Supplemental Table S4. List of genetic instrumental variables for AF in the Mendelian randomization analysis**

| **SNP** | **Chromosome** | **Position* (GRCh37)** | **Gene** | **Effect allele** | **Other allele** | **Effect allele frequency** | **Beta** | **Standard error** | **P-value** |
| --- | --- | --- | --- | --- | --- | --- | --- | --- | --- |
| rs284277 | 1 | 10790797 | CASZ1 | C | A | 0.38 | 0.039 | 0.007 | 1.2x10^-9^ |
| rs7529220 | 1 | 22282619 | HSPG2 | C | T | 0.85 | 0.058 | 0.010 | 2.0x10^-10^ |
| rs2885697 | 1 | 41544279 | SCMH1 | G | T | 0.35 | 0.039 | 0.007 | 2.9x10^-10^ |
| rs11590635 | 1 | 49309764 | AGBL4 | A | G | 0.02 | 0.148 | 0.024 | 4.1x10^-9^ |
| rs146518726 | 1 | 51535039 | MIR6500 | A | G | 0.03 | 0.157 | 0.020 | 8.3x10^-15^ |
| rs1545300 | 1 | 112464004 | KCND3 | C | T | 0.69 | 0.058 | 0.007 | 1.5x10^-14^ |
| rs4073778 | 1 | 116297758 | CASQ2 | A | C | 0.56 | 0.049 | 0.005 | 5.0x10^-13^ |
| rs10465885 | 1 | 147232740 | GJA5 | C | T | 0.47 | 0.039 | 0.007 | 1.4x10^-10^ |
| rs79187193 | 1 | 147255831 | GJA5 | G | A | 0.94 | 0.113 | 0.016 | 3.1x10^-14^ |
| rs6689306 | 1 | 154395946 | KCNN3 | A | G | 0.41 | 0.058 | 0.007 | 1.2x10^-18^ |
| rs4999127 | 1 | 154714006 | KCNN3 | A | G | 0.84 | 0.095 | 0.009 | 1.8x10^-23^ |
| rs11264280 | 1 | 154862952 | KCNN3 | T | C | 0.33 | 0.131 | 0.007 | 3.1x10^-79^ |
| rs72700114 | 1 | 170193825 | LINC01142 | C | G | 0.08 | 0.199 | 0.015 | 3.3x10^-54^ |
| rs72700118 | 1 | 170194823 | LINC01142 | A | C | 0.13 | 0.131 | 0.009 | 2.0x10^-36^ |
| rs577676 | 1 | 170587340 | LINC01142 | C | T | 0.56 | 0.068 | 0.007 | 2.4x10^-20^ |
| rs10753933 | 1 | 203026214 | PPFIA4 | T | G | 0.45 | 0.058 | 0.007 | 9.8x10^-20^ |
| rs4951258 | 1 | 205691316 | NUCKS1, SLC41A1 | A | G | 0.42 | 0.039 | 0.007 | 2.1x10^-8^ |
| rs7578393 | 2 | 26165528 | KIF3C | T | C | 0.80 | 0.058 | 0.007 | 2.4x10^-12^ |
| rs11125871 | 2 | 61470126 | USP34 | C | T | 0.61 | 0.039 | 0.005 | 6.4x10^-9^ |
| rs2540949 | 2 | 65284231 | CEP68 | A | T | 0.62 | 0.068 | 0.007 | 2.9x10^-22^ |
| rs6747542 | 2 | 70106832 | GMCL1, ANXA4 | T | C | 0.54 | 0.058 | 0.007 | 1.1x10^-16^ |
| rs72926475 | 2 | 86594487 | REEP1 | G | A | 0.88 | 0.068 | 0.010 | 2.4x10^-11^ |
| rs28387148 | 2 | 127433465 | GYPC | T | C | 0.11 | 0.077 | 0.012 | 6.3x10^-11^ |
| rs67969609 | 2 | 145760353 | TEX41 | G | C | 0.07 | 0.068 | 0.012 | 1.7x10^-8^ |
| rs56181519 | 2 | 175555714 | WIPF1 | C | T | 0.73 | 0.068 | 0.007 | 6.5x10^-18^ |
| rs2288327 | 2 | 179411665 | TTN, MIR548N, FKBP7, TTN-AS1 | G | A | 0.16 | 0.095 | 0.009 | 7.3x10^-25^ |
| rs3820888 | 2 | 201180023 | SPATS2L | C | T | 0.39 | 0.068 | 0.007 | 5.7x10^-24^ |
| rs35544454 | 2 | 213266003 | ERBB4 | A | T | 0.81 | 0.058 | 0.010 | 1.1x10^-11^ |
| rs7650482 | 3 | 12841804 | CAND2 | G | A | 0.64 | 0.068 | 0.007 | 1.8x10^-24^ |
| rs73041705 | 3 | 24463235 | THRB | T | C | 0.70 | 0.049 | 0.007 | 1.5x10^-9^ |
| rs7374540 | 3 | 38634142 | SCN10A, SCN5A | A | C | 0.61 | 0.049 | 0.007 | 7.4x10^-12^ |
| rs7373065 | 3 | 38710315 | SCN10A, SCN5A | T | C | 0.05 | 0.207 | 0.025 | 2.7x10^-16^ |
| rs6790396 | 3 | 38771925 | SCN10A, SCN5A | G | C | 0.60 | 0.058 | 0.007 | 2.4x10^-20^ |
| rs34080181 | 3 | 66454191 | LRIG1, SLC25A26 | G | A | 0.62 | 0.049 | 0.007 | 1.3x10^-10^ |
| rs17005647 | 3 | 69406181 | FRMD4B | T | C | 0.36 | 0.039 | 0.007 | 2.7x10^-9^ |
| rs6771054 | 3 | 89489529 | EPHA3 | T | C | 0.60 | 0.049 | 0.007 | 2.4x10^-11^ |
| rs10804493 | 3 | 111554426 | PHLDB2, PLCXD2 | A | G | 0.65 | 0.058 | 0.007 | 1.6x10^-15^ |
| rs1278493 | 3 | 135814009 | PPP2R3A | G | A | 0.44 | 0.039 | 0.005 | 8.8x10^-9^ |
| rs7612445 | 3 | 179172979 | GNB4 | T | G | 0.19 | 0.049 | 0.010 | 4.8x10^-9^ |
| rs60902112 | 3 | 194800853 | XXYLT1 | T | C | 0.23 | 0.049 | 0.007 | 1.7x10^-8^ |
| rs1458038 | 4 | 81164723 | FGF5 | T | C | 0.31 | 0.039 | 0.007 | 1.7x10^-9^ |
| rs10006327 | 4 | 103890980 | SLC9B1 | C | T | 0.49 | 0.039 | 0.007 | 4.4x10^-8^ |
| rs244017 | 4 | 111255917 | PITX2 | T | G | 0.78 | 0.058 | 0.010 | 7.7x10^-11^ |
| rs61501369 | 4 | 111524629 | PITX2 | T | C | 0.23 | 0.095 | 0.007 | 1.0x10^-28^ |
| rs6850025 | 4 | 111596360 | PITX2 | A | G | 0.05 | 0.113 | 0.018 | 9.5x10^-11^ |
| rs67249485 | 4 | 111699685 | PITX2 | T | A | 0.20 | 0.365 | 0.007 | 7.3x10^-443^ |
| rs3853445 | 4 | 111761487 | PITX2 | T | C | 0.73 | 0.148 | 0.011 | 3.7x10^-52^ |
| rs79399769 | 4 | 111925656 | PITX2 | C | T | 0.97 | 0.215 | 0.025 | 2.0x10^-18^ |
| rs1532170 | 4 | 112165212 | PITX2 | G | A | 0.45 | 0.068 | 0.007 | 8.0x10^-18^ |
| rs138311480 | 4 | 112454295 | PITX2 | C | T | 0.98 | 0.166 | 0.028 | 3.0x10^-9^ |
| rs114904067 | 4 | 112604821 | PITX2 | G | A | 0.97 | 0.122 | 0.020 | 8.6x10^-9^ |
| rs7687819 | 4 | 113329345 | PITX2 | A | G | 0.77 | 0.049 | 0.007 | 8.7x10^-9^ |
| rs6829664 | 4 | 114448656 | CAMK2D | G | A | 0.26 | 0.058 | 0.007 | 1.9x10^-13^ |
| rs10213171 | 4 | 148937537 | ARHGAP10 | G | C | 0.06 | 0.095 | 0.012 | 1.3x10^-11^ |
| rs10520260 | 4 | 174447349 | HAND2, HAND2-AS1 | A | G | 0.68 | 0.049 | 0.007 | 5.9x10^-10^ |
| rs12648245 | 4 | 174641184 | HAND2, HAND2-AS1 | T | C | 0.92 | 0.095 | 0.012 | 3.5x10^-13^ |
| rs6596717 | 5 | 106427609 | LOC102467213 | C | A | 0.40 | 0.039 | 0.007 | 3.0x10^-9^ |
| rs337705 | 5 | 113737062 | KCNN2 | G | T | 0.38 | 0.058 | 0.007 | 1.6x10^-16^ |
| rs2012809 | 5 | 128190363 | SLC27A6 | G | A | 0.79 | 0.058 | 0.010 | 4.9x10^-10^ |
| rs2040862 | 5 | 137419989 | WNT8A, NPY6R, MYOT, FAM13B | T | C | 0.18 | 0.104 | 0.007 | 1.1x10^-35^ |
| rs6580277 | 5 | 142818123 | NR3C1 | G | A | 0.24 | 0.068 | 0.010 | 1.6x10^-17^ |
| rs12188351 | 5 | 168386089 | SLIT3 | A | G | 0.06 | 0.086 | 0.014 | 2.5x10^-9^ |
| rs6891790 | 5 | 172670745 | NKX2-5 | G | T | 0.72 | 0.077 | 0.007 | 4.5x10^-22^ |
| rs28439930 | 5 | 173393111 | NKX2-5 | G | C | 0.52 | 0.049 | 0.007 | 1.9x10^-11^ |
| rs73366713 | 6 | 16415751 | ATXN1 | G | A | 0.86 | 0.104 | 0.009 | 1.5x10^-25^ |
| rs34969716 | 6 | 18210109 | KDM1B, DEK | A | G | 0.31 | 0.068 | 0.007 | 1.6x10^-19^ |
| rs3176326 | 6 | 36647289 | CDKN1A, PANDAR, PI16 | G | A | 0.80 | 0.058 | 0.007 | 1.4x10^-13^ |
| rs2031522 | 6 | 87821501 | CGA | A | G | 0.62 | 0.039 | 0.007 | 1.5x10^-10^ |
| rs3951016 | 6 | 118559658 | SLC35F1, PLN | A | T | 0.46 | 0.068 | 0.007 | 2.1x10^-22^ |
| rs9401451 | 6 | 122099152 | HSF2 | G | A | 0.90 | 0.077 | 0.012 | 3.4x10^-11^ |
| rs13195459 | 6 | 122403559 | HSF2 | G | A | 0.64 | 0.058 | 0.007 | 4.2x10^-19^ |
| rs117984853 | 6 | 149399100 | UST | T | G | 0.10 | 0.122 | 0.014 | 1.3x10^-24^ |
| rs55734480 | 7 | 14372009 | DGKB | A | G | 0.25 | 0.058 | 0.007 | 2.2x10^-12^ |
| rs6462079 | 7 | 28415827 | CREB5 | A | G | 0.72 | 0.049 | 0.007 | 8.8x10^-10^ |
| rs35005436 | 7 | 74134911 | GTF2I, LOC101926943, GTF2IRD2 | C | T | 0.16 | 0.058 | 0.010 | 3.3x10^-10^ |
| rs56201652 | 7 | 92278116 | CDK6 | G | A | 0.73 | 0.049 | 0.007 | 1.7x10^-12^ |
| rs11773845 | 7 | 116191301 | CAV1, CAV2 | A | C | 0.59 | 0.104 | 0.007 | 2.4x10^-55^ |
| rs55985730 | 7 | 128417044 | OPN1SW, CALU | G | T | 0.06 | 0.086 | 0.014 | 5.2x10^-9^ |
| rs7789146 | 7 | 150661409 | KCNH2 | G | A | 0.82 | 0.058 | 0.010 | 2.1x10^-11^ |
| rs35620480 | 8 | 11499908 | GATA4 | C | A | 0.16 | 0.058 | 0.007 | 5.2x10^-9^ |
| rs7508 | 8 | 17913970 | ASAH1 | A | G | 0.71 | 0.068 | 0.007 | 1.7x10^-21^ |
| rs7834729 | 8 | 21821778 | XPO7 | G | T | 0.89 | 0.068 | 0.010 | 3.6x10^-10^ |
| rs62521286 | 8 | 124551975 | FBXO32 | G | A | 0.07 | 0.122 | 0.014 | 4.5x10^-19^ |
| rs4871397 | 8 | 124635197 | FBXO32 | G | C | 0.09 | 0.086 | 0.014 | 1.3x10^-9^ |
| rs6994744 | 8 | 141740868 | PTK2 | C | A | 0.50 | 0.039 | 0.005 | 1.1x10^-9^ |
| rs10821415 | 9 | 97713459 | C9orf3 | A | C | 0.41 | 0.086 | 0.007 | 2.9x10^-34^ |
| rs2274115 | 9 | 139094773 | LHX3 | G | A | 0.70 | 0.049 | 0.010 | 1.7x10^-10^ |
| rs12245149 | 10 | 65321147 | REEP3, NRBF2 | C | A | 0.53 | 0.049 | 0.007 | 1.7x10^-12^ |
| rs7096385 | 10 | 69664881 | SIRT1, MYPN | T | C | 0.09 | 0.068 | 0.012 | 4.9x10^-8^ |
| rs60212594 | 10 | 75414344 | SYNPO2L, NUDT13, MYOZ1, AGAP5 | G | C | 0.86 | 0.113 | 0.011 | 9.2x10^-35^ |
| rs10458660 | 10 | 77936576 | C10orf11 | G | A | 0.17 | 0.058 | 0.007 | 6.8x10^-10^ |
| rs55693294 | 10 | 105277474 | NEURL1 | T | C | 0.06 | 0.086 | 0.014 | 4.2x10^-9^ |
| rs11598047 | 10 | 105342672 | NEURL1 | G | A | 0.16 | 0.157 | 0.009 | 9.0x10^-66^ |
| rs35176054 | 10 | 105480387 | NEURL1 | A | T | 0.13 | 0.140 | 0.011 | 8.2x10^-41^ |
| rs10749053 | 10 | 112576695 | RBM20 | T | C | 0.16 | 0.058 | 0.010 | 1.0x10^-8^ |
| rs10741807 | 11 | 20011445 | NAV2 | T | C | 0.25 | 0.077 | 0.007 | 1.6x10^-20^ |
| rs4935786 | 11 | 121661507 | SORL1 | T | A | 0.27 | 0.049 | 0.007 | 4.9x10^-9^ |
| rs76097649 | 11 | 128764570 | KCNJ5 | A | G | 0.09 | 0.113 | 0.011 | 1.3x10^-20^ |
| rs2291437 | 12 | 24715048 | LINC00477 | G | T | 0.11 | 0.086 | 0.012 | 2.1x10^-17^ |
| rs4963776 | 12 | 24779491 | LINC00477 | G | T | 0.82 | 0.095 | 0.007 | 1.8x10^-25^ |
| rs17380837 | 12 | 26345526 | SSPN | C | T | 0.69 | 0.049 | 0.007 | 4.8x10^-12^ |
| rs12809354 | 12 | 32978437 | PKP2 | C | T | 0.14 | 0.068 | 0.010 | 2.9x10^-14^ |
| rs11614818 | 12 | 56055815 | NACA | C | T | 0.36 | 0.039 | 0.005 | 1.9x10^-8^ |
| rs2860482 | 12 | 57105938 | NACA | A | C | 0.27 | 0.058 | 0.007 | 1.2x10^-12^ |
| rs71454237 | 12 | 70013415 | LRRC10 | G | A | 0.79 | 0.058 | 0.007 | 1.8x10^-13^ |
| rs775498 | 12 | 70071513 | LRRC10 | G | A | 0.28 | 0.039 | 0.007 | 9.4x10^-9^ |
| rs12426679 | 12 | 76237987 | PHLDA1 | C | T | 0.47 | 0.039 | 0.005 | 4.9x10^-9^ |
| rs883079 | 12 | 114793240 | TBX5 | T | C | 0.71 | 0.095 | 0.007 | 2.8x10^-40^ |
| rs10773657 | 12 | 123327900 | HIP1R | C | A | 0.14 | 0.058 | 0.010 | 2.5x10^-8^ |
| rs6560886 | 12 | 133150210 | FBRSL1 | C | T | 0.79 | 0.049 | 0.010 | 1.5x10^-8^ |
| rs9506925 | 13 | 23368943 | LINC00540, LINC00621, SGCG | T | C | 0.27 | 0.049 | 0.007 | 2.7x10^-9^ |
| rs35569628 | 13 | 113872712 | CUL4A | T | C | 0.78 | 0.049 | 0.007 | 1.4x10^-8^ |
| rs422068 | 14 | 23864804 | MYH6, MYH7 | C | T | 0.35 | 0.039 | 0.007 | 3.9x10^-10^ |
| rs1957021 | 14 | 32924505 | AKAP6 | C | T | 0.22 | 0.058 | 0.007 | 4.8x10^-15^ |
| rs11156751 | 14 | 32990437 | AKAP6 | C | T | 0.29 | 0.068 | 0.007 | 6.9x10^-21^ |
| rs73241997 | 14 | 35173775 | CFL2 | T | C | 0.14 | 0.077 | 0.009 | 2.9x10^-15^ |
| rs2738413 | 14 | 64679960 | SYNE2, MIR548AZ, ESR2, MTHFD1 | A | G | 0.50 | 0.077 | 0.007 | 2.5x10^-31^ |
| rs74884082 | 14 | 73249419 | DPF3 | C | T | 0.75 | 0.049 | 0.010 | 3.5x10^-10^ |
| rs10873298 | 14 | 77426525 | IRF2BPL | C | T | 0.37 | 0.039 | 0.007 | 7.1x10^-9^ |
| rs147301839 | 15 | 57924714 | GCOM1/MYZAP | C | A | 0.01 | 0.329 | 0.053 | 1.9x10^-10^ |
| rs7170477 | 15 | 64103777 | HERC1 | A | G | 0.30 | 0.039 | 0.005 | 5.0x10^-8^ |
| rs74022964 | 15 | 73677264 | HCN4 | T | C | 0.16 | 0.113 | 0.009 | 3.5x10^-36^ |
| rs12908004 | 15 | 80676925 | ARNT2 | G | A | 0.16 | 0.077 | 0.009 | 4.1x10^-16^ |
| rs2759301 | 15 | 80994288 | ARNT2 | A | G | 0.45 | 0.039 | 0.005 | 9.2x10^-9^ |
| rs4965430 | 15 | 99268850 | IGF1R | C | G | 0.39 | 0.049 | 0.007 | 1.3x10^-10^ |
| rs118159104 | 16 | 1676804 | RPL3L | G | T | 0.01 | 0.182 | 0.032 | 1.6x10^-8^ |
| rs140185678 | 16 | 2003016 | RPL3L | A | G | 0.04 | 0.166 | 0.022 | 2.4x10^-14^ |
| rs77316573 | 16 | 2265271 | RPL3L | T | C | 0.20 | 0.049 | 0.010 | 2.4x10^-8^ |
| rs2359171 | 16 | 73053022 | ZFHX3 | A | T | 0.18 | 0.174 | 0.009 | 4.6x10^-91^ |
| rs876727 | 16 | 73067761 | ZFHX3 | T | G | 0.79 | 0.049 | 0.010 | 6.8x10^-9^ |
| rs7225165 | 17 | 1309850 | YWHAE, CRK, MYO1C | G | A | 0.89 | 0.068 | 0.012 | 3.2x10^-9^ |
| rs9899183 | 17 | 7452977 | TNFSF12, TNFSF12-TNFSF13, SOX15, FXR2 | T | C | 0.71 | 0.049 | 0.007 | 2.0x10^-9^ |
| rs72811294 | 17 | 12618680 | MYOCD | G | C | 0.89 | 0.068 | 0.012 | 9.7x10^-12^ |
| rs11658278 | 17 | 38031164 | ZPBP2, GSDMB, ORMDL3 | T | C | 0.48 | 0.049 | 0.007 | 3.5x10^-11^ |
| rs1563304 | 17 | 44874453 | WNT3 | T | C | 0.18 | 0.068 | 0.010 | 2.6x10^-12^ |
| rs12604076 | 17 | 76773638 | CYTH1, USP36 | T | C | 0.48 | 0.039 | 0.007 | 3.6x10^-8^ |
| rs9953366 | 18 | 46474192 | SMAD7 | C | T | 0.66 | 0.049 | 0.007 | 1.8x10^-11^ |
| rs9963878 | 18 | 48679522 | MEX3C | C | T | 0.09 | 0.068 | 0.012 | 2.5x10^-8^ |
| rs8088085 | 18 | 48708548 | MEX3C | A | C | 0.54 | 0.039 | 0.007 | 4.8x10^-8^ |
| rs2834618 | 21 | 36119111 | LINC01426 | T | G | 0.89 | 0.095 | 0.009 | 3.4x10^-17^ |
| rs464901 | 22 | 18597502 | TUBA8 | T | C | 0.67 | 0.049 | 0.007 | 1.5x10^-12^ |
| rs133902 | 22 | 26164079 | MYO18B | T | C | 0.43 | 0.039 | 0.007 | 9.1x10^-10^ |

* Genomic positions for all listed variants are reported based on the GRCh37/hg19 reference build.

Abbreviations: AF, atrial fibrillation; GRCh37, Genome Reference Consortium human genome build 37; hg19, human genome version 19; SNP, single nucleotide polymorphism.

**Supplemental Table S5. List of genetic instrumental variables for HF in the Mendelian randomization analysis**

| **SNP** | **Chromosome** | **Position* (GRCh37)** | **Gene** | **Effect allele** | **Other allele** | **Effect allele frequency** | **Beta** | **Standard error** | **P-value** |
| --- | --- | --- | --- | --- | --- | --- | --- | --- | --- |
| rs660240 | 1 | 109817838 | CELSR2 | C | T | 0.79 | 0.058 | 0.010 | 3.3x10^-10^ |
| rs17042102 | 4 | 111668626 | PITX2, FAM241A | A | G | 0.12 | 0.113 | 0.011 | 5.7x10^-20^ |
| rs11745324 | 5 | 137012171 | KLHL3 | G | A | 0.77 | 0.049 | 0.010 | 2.4x10^-8^ |
| rs4135240 | 6 | 36647680 | CDKN1A | T | C | 0.66 | 0.049 | 0.010 | 6.8x10^-9^ |
| rs55730499 | 6 | 161005610 | LPA | T | C | 0.07 | 0.104 | 0.014 | 1.8x10^-11^ |
| rs140570886 | 6 | 161013013 | LPA | C | T | 0.02 | 0.215 | 0.029 | 7.7x10^-11^ |
| rs1556516 | 9 | 22100176 | 9p21/CDKN2B-AS1 | C | G | 0.48 | 0.058 | 0.007 | 1.6x10^-15^ |
| rs600038 | 9 | 136151806 | ABO, SURF1 | C | T | 0.21 | 0.058 | 0.010 | 3.7x10^-9^ |
| rs4746140 | 10 | 75417249 | SYNPO2L, AGAP5 | G | C | 0.85 | 0.068 | 0.010 | 1.1x10^-9^ |
| rs17617337 | 10 | 121426884 | BAG3 | C | T | 0.78 | 0.058 | 0.010 | 3.7x10^-9^ |
| rs4766578 | 12 | 111904371 | ATXN2 | T | A | 0.47 | 0.039 | 0.007 | 4.9x10^-8^ |
| rs56094641 | 16 | 53806453 | FTO | G | A | 0.42 | 0.049 | 0.007 | 1.2x10^-8^ |

* Genomic positions for all listed variants are reported based on the GRCh37/hg19 reference build.

Abbreviations: GRCh37, Genome Reference Consortium human genome build 37; HF, heart failure; hg19, human genome version 19; SNP, single nucleotide polymorphism.

**Supplemental Table S6. Comparisons of CMR imaging measurements by AI-ECG age gap quartiles**

| **Measurements of CMR** | **AI-ECG age gap quartiles** | | | | | **P-value** |
| --- | --- | --- | --- | --- | --- | --- |
|  | **Total**  **(N = 35090)** | **Quartile 1**  **(N = 8773)** | **Quartile 2**  **(N = 8773)** | **Quartile 3**  **(N = 8772)** | **Quartile 4**  **(N = 8772)** |  |
| **LA parameters** |  |  |  |  |  |  |
| LA maximum volume (mL) | 72.2 ± 22.5 | 69.7 ± 21.4 | 71.2 ± 22.4 | 73.0 ± 23.3 | 74.7 ± 22.7 | <0.001 |
| Indexed LA maximum volume (mL/m^2^) | 38.8 ± 11.0 | 38.0 ± 10.9 | 38.5 ± 11.1 | 39.0 ± 11.2 | 39.5 ± 10.9 | <0.001 |
| LA minimum volume (mL) | 28.7 ± 13.9 | 27.8 ± 12.7 | 28.3 ± 13.8 | 29.0 ± 14.8 | 29.6 ± 14.1 | <0.001 |
| Indexed LA minimum volume (mL/m^2^) | 15.4 ± 7.0 | 15.2 ± 6.6 | 15.3 ± 7.0 | 15.5 ± 7.3 | 15.6 ± 6.9 | 0.002 |
| LA stroke volume (mL) | 43.5 ± 11.8 | 41.8 ± 11.3 | 42.9 ± 11.7 | 44.0 ± 11.9 | 45.1 ± 12.0 | <0.001 |
| Indexed LA stroke volume (mL/m^2^) | 23.4 ± 5.8 | 22.8 ± 5.8 | 23.3 ± 5.8 | 23.6 ± 5.8 | 23.9 ± 5.8 | <0.001 |
| LA ejection fraction (%) | 61.6 ± 8.9 | 61.3 ± 8.8 | 61.6 ± 8.9 | 61.7 ± 9.0 | 61.8 ± 9.0 | 0.011 |
| **LV parameters** |  |  |  |  |  |  |
| LV end-diastolic volume (mL) | 147.9 ± 33.7 | 142.6 ± 32.3 | 146.4 ± 33.1 | 149.5 ± 34.3 | 153.1 ± 34.0 | <0.001 |
| Indexed LV end-diastolic volume (mL/m^2^) | 79.3 ± 14.0 | 77.5 ± 13.8 | 79.0 ± 13.8 | 79.8 ± 14.3 | 80.7 ± 13.8 | <0.001 |
| LV end-systolic volume (mL) | 60.3 ± 19.1 | 58.0 ± 18.1 | 59.5 ± 18.6 | 61.0 ± 20.0 | 62.7 ± 19.3 | <0.001 |
| Indexed LV end-systolic volume (mL/m^2^) | 32.2 ± 8.6 | 31.4 ± 8.3 | 32.0 ± 8.5 | 32.5 ± 9.1 | 32.9 ± 8.5 | <0.001 |
| LV stroke volume (mL) | 87.6 ± 19.2 | 84.6 ± 18.5 | 86.8 ± 18.9 | 88.5 ± 19.1 | 90.5 ± 19.6 | <0.001 |
| Indexed LV stroke volume (mL/m^2^) | 47.1 ± 8.4 | 46.1 ± 8.4 | 46.9 ± 8.3 | 47.3 ± 8.3 | 47.8 ± 8.5 | <0.001 |
| LV ejection fraction (%) | 59.6 ± 6.0 | 59.7 ± 6.0 | 59.7 ± 6.0 | 59.6 ± 6.0 | 59.4 ± 6.1 | 0.019 |
| LV longitudinal strain (%) | -18.5 ± 2.8 | -18.6 ± 2.7 | -18.6 ± 2.7 | -18.6 ± 2.8 | -18.5 ± 2.8 | 0.07 |
| LV circumferential strain (%) | -22.3 ± 3.3 | -22.5 ± 3.2 | -22.4 ± 3.3 | -22.3 ± 3.3 | -22.1 ± 3.4 | <0.001 |
| LV radial strain (%) | 45.2 ± 8.2 | 45.5 ± 8.1 | 45.3 ± 8.1 | 45.2 ± 8.3 | 44.8 ± 8.4 | <0.001 |
| Myocardial mass (g) | 86.0 ± 22.2 | 83.5 ± 21.1 | 85.1 ± 21.8 | 86.7 ± 22.6 | 88.6 ± 22.7 | <0.001 |
| Indexed myocardial mass (g/m^2^) | 45.8 ± 8.5 | 45.1 ± 8.2 | 45.7 ± 8.4 | 45.9 ± 8.7 | 46.4 ± 8.6 | <0.001 |
| LV mean wall thickness (mm) | 5.7 ± 0.8 | 5.7 ± 0.8 | 5.7 ± 0.8 | 5.7 ± 0.8 | 5.7 ± 0.8 | <0.001 |
| Indexed LV mean wall thickness (mm/m^2^) | 3.1 ± 0.3 | 3.1 ± 0.3 | 3.1 ± 0.3 | 3.1 ± 0.3 | 3.0 ± 0.3 | <0.001 |
| **Other parameters** |  |  |  |  |  |  |
| Pericardial adipose tissue area (cm^2^) | 23.5 ± 12.8 | 23.2 ± 12.7 | 23.2 ± 12.8 | 23.6 ± 12.8 | 24.0 ± 13.0 | <0.001 |
| Indexed pericardial adipose tissue area (cm²/m²) | 12.4 ± 6.0 | 12.4 ± 6.0 | 12.2 ± 6.0 | 12.3 ± 6.0 | 12.4 ± 6.0 | 0.17 |

All CMR measurements are presented as mean ± standard deviation. Indexed values represent measurements normalized to body surface area. P-values for comparisons across quartile groups were calculated using one-way analysis of variance (ANOVA) or the Kruskal–Wallis test, where appropriate.

Abbreviations: AI, artificial intelligence; CMR, cardiac magnetic resonance; ECG, electrocardiogram; LA, left atrium; LV, left ventricle.

**Supplemental Table S7. Incidence and risk of AF across AI-ECG age gap quartiles**

| **AI-ECG age gap quartiles** | **No. of events / total No. (%)** | **Event rates***  **(95% CI)** | **Model 1**† | | **Model 2**‡ | |
| --- | --- | --- | --- | --- | --- | --- |
|  |  |  | **HR (95% CI)** | **P-value** | **HR (95% CI)** | **P-value** |
| Quartile 1 | 117 / 8773 (1.33%) | 3.93 (3.24–4.77) | 1.00 [ref] | – [ref] | 1.00 [ref] | – [ref] |
| Quartile 2 | 120 / 8773 (1.37%) | 4.70 (3.93–5.62) | 1.31 (1.02–1.69) | 0.038 | 1.27 (0.98–1.64) | 0.06 |
| Quartile 3 | 119 / 8772 (1.36%) | 6.17 (5.09–7.46) | 1.59 (1.23–2.07) | <0.001 | 1.51 (1.16–1.96) | 0.002 |
| Quartile 4 | 132 / 8772 (1.50%) | 10.70 (8.36–13.70) | 2.59 (1.96–3.41) | <0.001 | 2.38 (1.80–3.14) | <0.001 |

* The event rates were adjusted for chronological age and sex and presented per 1,000 person-years.

† Model 1 was adjusted for chronological age and sex.

‡ Model 2 was adjusted for chronological age, sex, Townsend deprivation index, smoking status, alcohol consumption, physical activity levels, BMI, systolic BP, hemoglobin A1c, LDL cholesterol, and eGFR.

Abbreviations: AF, atrial fibrillation; AI, artificial intelligence; BMI, body mass index; BP, blood pressure; CI, confidence interval; ECG, electrocardiogram; eGFR, estimated glomerular filtration rate; HR, hazard ratio; LDL, low-density lipoprotein.

**Supplemental Table S8. Risk of AF across AI-ECG age gap quartiles accounting for competing risk of death**

| **AI-ECG age gap quartiles** | **Model 1*** | | | **Model 2**† | |
| --- | --- | --- | --- | --- | --- |
|  | **HR (95% CI)** | **P-value** | **HR (95% CI)** | | **P-value** |
| Quartile 1 | 1.00 [ref] | – [ref] | 1.00 [ref] | | – [ref] |
| Quartile 2 | 1.13 (1.01–1.70) | 0.039 | 1.27 (0.98–1.65) | | 0.07 |
| Quartile 3 | 1.59 (1.22–2.08) | 0.001 | 1.51 (1.15–1.98) | | 0.003 |
| Quartile 4 | 2.58 (1.04–3.43) | <0.001 | 2.38 (1.78–3.18) | | <0.001 |

† Model 1 was adjusted for chronological age and sex.

‡ Model 2 was adjusted for chronological age, sex, Townsend deprivation index, smoking status, alcohol consumption, physical activity levels, BMI, systolic BP, hemoglobin A1c, LDL cholesterol, and eGFR.

Abbreviations: AF, atrial fibrillation; AI, artificial intelligence; BMI, body mass index; BP, blood pressure; CI, confidence interval; ECG, electrocardiogram; eGFR, estimated glomerular filtration rate; HR, hazard ratio; LDL, low-density lipoprotein.

**Supplemental Table S9. Heterogeneity and horizontal pleiotropy assessments for Mendelian randomization analyses of ECG-aging and AF**

| **Exposure** | **Outcome** | **Outcome dataset (Population)** | **No. of SNPs** | **Heterogeneity (IVW)** | | **MR-Egger** | | **MR-PRESSO** | | |
| --- | --- | --- | --- | --- | --- | --- | --- | --- | --- | --- |
|  |  |  |  | **Cochran’s Q** | **P-value** | **Intercept (SE)** | **P-value** | **Global P-value** | **No. of outliers** | **Distortion P-value** |
| **ECG-aging** | **AF** | Nielsen et al.  (European) | 8 | 193.69 | <0.001 | -0.072 (0.046) | 0.17 | 0.23 | 0 | NA |
|  |  | FinnGen study  (European) | 8 | 109.68 | <0.001 | -0.055 (0.047) | 0.28 | <0.001 | 3 | 0.76 |
| **AF** | **ECG-aging** | UK Biobank | 108 | 331.50 | <0.001 | 0.030 (0.018) | 0.10 | <0.001 | 8 | 0.69 |

Cochran’s Q statistics from IVW models were used to assess heterogeneity among SNP-specific causal estimates. The MR-Egger intercept test evaluated directional horizontal pleiotropy. The MR-PRESSO global test assessed the presence of outlier-driven pleiotropy, and distortion tests compared causal estimates before and after outlier removal when applicable.

Abbreviations: AF, atrial fibrillation; ECG-aging, electrocardiographic aging; IVW, inverse-variance weighted; MR, Mendelian randomization; PRESSO, pleiotropy residual sum and outlier; SE, standard error; SNP, single-nucleotide polymorphism.

**Supplemental Table S10. Steiger-filtered bidirectional Mendelian randomization results of the causal association between ECG-aging and AF risk**

| **Exposure** | **Outcome** | **Methods** | **Effect size**  **(95% CI)** | **P-value** |
| --- | --- | --- | --- | --- |
| ECG-aging | AF | MR-IVW | 1.13 (1.02–1.25) | 0.025 |
|  |  | MR-Egger | 1.40 (1.05–1.87) | 0.032 |
|  |  | Weighted median | 1.06 (1.01–1.11) | 0.011 |
|  |  | MR-RAPS | 1.20 (1.17–1.24) | <0.001 |
| AF | ECG-aging | MR-IVW | 1.02 (0.87–1.20) | 0.79 |
|  |  | MR-Egger | 0.87 (0.60–1.28) | 0.48 |
|  |  | Weighted median | 0.97 (0.78–1.21) | 0.78 |
|  |  | MR-RAPS | 1.02 (0.90–1.17 | 0.75 |

GWAS summary data for ECG-aging and AF (Nielsen et al.) were alternately used as exposure and outcome in the bidirectional framework. MR estimates for ECG-aging to AF were identical to those of the main analyses, since no SNPs were excluded after applying Steiger filtering.

Abbreviations: AF, atrial fibrillation; CI, confidence interval; ECG-aging, electrocardiographic aging; GWAS, genome-wide association study; IVW, inverse-variance weighted; MR, Mendelian randomization; PRESSO, pleiotropy residual sum and outlier; RAPS, robust adjusted profile score.

**Supplemental Table S11. Multivariable regression analyses of the associations between AI-ECG age gap and CMR imaging measurements**

| **Measurements of CMR** | **Model 1*** | | **Model 2†** | |
| --- | --- | --- | --- | --- |
|  | **OR (95% CI)** | **P-value** | **OR (95% CI)** | **P-value** |
| **LA parameters** |  |  |  |  |
| LA maximum volume (mL) | 3.50 (2.65–4.63) | <0.001 | 2.93 (2.16–3.97) | <0.001 |
| LA minimum volume (mL) | 3.09 (2.55–3.74) | <0.001 | 2.98 (2.41–3.67) | <0.001 |
| LA stroke volume (mL) | 1.13 (0.98–1.30) | 0.08 | 0.98 (0.85–1.15) | 0.84 |
| LA ejection fraction (%) | 0.67 (0.60–0.76) | <0.001 | 0.64 (0.56–0.73) | <0.001 |
| **LV parameters** |  |  |  |  |
| LV end-diastolic volume (mL) | 2.03 (1.49–2.77) | <0.001 | 1.44 (1.03–2.02) | 0.034 |
| LV end-systolic volume (mL) | 1.48 (1.22–1.80) | <0.001 | 1.42 (1.15–1.76) | 0.001 |
| LV stroke volume (mL) | 1.37 (1.13–1.67) | 0.002 | 1.01 (0.82–1.26) | 0.90 |
| LV ejection fraction (%) | 0.99 (0.92–1.07) | 0.89 | 0.93 (0.86–1.01) | 0.09 |
| LV longitudinal strain (%) | 0.95 (0.91–0.98) | 0.002 | 0.94 (0.90–0.98) | 0.002 |
| LV circumferential strain (%) | 0.91 (0.87–0.95) | <0.001 | 0.88 (0.84–0.92) | <0.001 |
| LV radial strain (%) | 1.23 (1.11–1.36) | <0.001 | 1.05 (0.95–1.17) | 0.35 |
| Myocardial mass (g) | 1.93 (1.63–2.28) | <0.001 | 1.26 (1.05–1.50) | 0.012 |
| LV mean wall thickness (mm) | 1.03 (1.03–1.04) | <0.001 | 1.02 (1.01–1.02) | <0.001 |
| **Other parameters** |  |  |  |  |
| Pericardial adipose tissue area (cm^2^) | 1.60 (1.41–1.82) | <0.001 | 1.57 (1.36–1.80) | <0.001 |

Estimates are reported as adjusted ORs per 1-SD increment in the AI-ECG age gap, corresponding to each CMR parameter.

* Model 1 was adjusted for chronological age, sex, and body mass index.

**†** Model 2 was further adjusted for Townsend deprivation index, smoking status, alcohol consumption, systolic blood pressure, hemoglobin A1c, low-density lipoprotein cholesterol, and estimated glomerular filtration rate.

Abbreviations: AI, artificial intelligence; CMR, cardiac magnetic resonance; ECG, electrocardiogram; LA, left atrium; LV, left ventricle; OR, odds ratio; SD, standard deviation.

**Supplemental Table S12. GRS-based Mendelian randomization analyses of associations between genetically predicted ECG-aging and CMR imaging measurements**

| **Measurements of CMR** | **Model 1*** | | **Model 2†** | |
| --- | --- | --- | --- | --- |
|  | **OR (95% CI)** | **P-value** | **OR (95% CI)** | **P-value** |
| **LA parameters** |  |  |  |  |
| LA maximum volume (mL) | 1.06 (0.80–1.40) | 0.70 | 1.11 (0.82–1.50) | 0.51 |
| LA minimum volume (mL) | 1.08 (0.89–1.31) | 0.44 | 1.12 (0.91–1.38) | 0.27 |
| LA stroke volume (mL) | 0.98 (0.85–1.13) | 0.78 | 0.99 (0.85–1.14) | 0.85 |
| LA ejection fraction (%) | 0.96 (0.85–1.08) | 0.50 | 0.94 (0.82–1.06) | 0.31 |
| **LV parameters** |  |  |  |  |
| LV end-diastolic volume (mL) | 0.79 (0.58–1.08) | 0.14 | 0.90 (0.64–1.25) | 0.51 |
| LV end-systolic volume (mL) | 0.82 (0.68–0.99) | 0.043 | 0.89 (0.73–1.10) | 0.27 |
| LV stroke volume (mL) | 0.96 (0.79–1.17) | 0.72 | 1.00 (0.81–1.24) | 0.97 |
| LV ejection fraction (%) | 1.05 (0.98–1.13) | 0.17 | 1.03 (0.95–1.12) | 0.44 |
| LV longitudinal strain (%) | 1.03 (0.99–1.06) | 0.12 | 1.02 (0.99–1.06) | 0.21 |
| LV circumferential strain (%) | 0.99 (0.95–1.03) | 0.59 | 1.00 (0.96–1.04) | 0.94 |
| LV radial strain (%) | 1.04 (0.94–1.15) | 0.43 | 1.01 (0.91–1.12) | 0.89 |
| Myocardial mass (g) | 1.26 (1.07–1.49) | 0.006 | 1.24 (1.05–1.48) | 0.014 |
| LV mean wall thickness (mm) | 1.01 (1.00–1.02) | 0.002 | 1.01 (1.01–1.02) | <0.001 |
| **Other parameters** |  |  |  |  |
| Pericardial adipose tissue area (cm^2^) | 1.01 (0.89–1.14) | 0.89 | 1.01 (0.88–1.16) | 0.89 |

Estimates are reported as adjusted ORs per 1-SD increment in the GRS for AI-ECG age gap, corresponding to each CMR parameter.

* Model 1 was adjusted for chronological age, sex, body mass index, genotype batch, assessment center, and 10 genetic principal components.

**†** Model 2 was further adjusted for Townsend deprivation index, smoking status, alcohol consumption, systolic blood pressure, hemoglobin A1c, low-density lipoprotein cholesterol, and estimated glomerular filtration rate.

Abbreviations: AI, artificial intelligence; CMR, cardiac magnetic resonance; ECG, electrocardiogram; ECG-aging, electrocardiographic aging; GRS, genetic risk score; LA, left atrium; LV, left ventricle; OR, odds ratio; SD, standard deviation.

**Supplemental Table S13. Two-sample Mendelian randomization analyses of the causal association between ECG-aging and AF using UK Biobank–excluded outcome GWAS summary statistics**

| **Exposure** | **Outcome** | **Outcome dataset** | **Number**  **of cases**  **/ Participants** | **Cochran’s Q statistics**  **P-value** | **MR-Egger**  **Intercept**  **P-value** | **MR Methods** | **Effect size**  **(95% CI)** | **P-value** |
| --- | --- | --- | --- | --- | --- | --- | --- | --- |
| ECG-aging | Atrial fibrillation | Nielsen et al.  (UK Biobank–excluded dataset) | 45,800  / 635,097 | <0.001 | 0.28 | MR-IVW | 1.13 (1.02–1.26) | 0.021 |
|  |  |  |  |  |  | MR-Egger | 1.36 (0.99–1.87) | 0.06 |
|  |  |  |  |  |  | Weighted median | 1.06 (1.01–1.12) | 0.030 |
|  |  |  |  |  |  | MR-RAPS | 1.19 (1.16–1.23) | <0.001 |
|  |  |  |  |  |  | MR-PRESSO* | 1.13 (1.02–1.26) | 0.021 |

Effect estimates were derived from two-sample MR using outcome GWAS data excluding UK Biobank participants, with complementary sensitivity MR results provided.

* The MR-PRESSO global test (Global p-value = 0.25) did not detect any significant outliers in the genetic instrument, and no correction was necessary. As a result, the causal estimate calculated by MR-PRESSO was the same as that by the IVW method.

Abbreviations: AF, atrial fibrillation; CI, confidence interval; ECG-aging, electrocardiographic aging; IVW, inverse-variance weighted; MR, Mendelian randomization; PRESSO, pleiotropy residual sum and outlier; GWAS, genome-wide association study; RAPS, robust adjusted profile score.

**Supplemental Figure S1. Schematics of Mendelian randomization analyses performed in this study**

**(A) Conceptual diagram illustrating assumptions of two-sample MR**

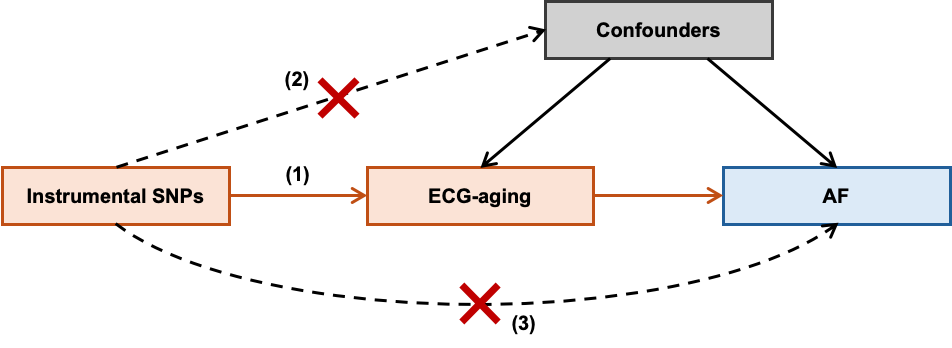

MR relies on the following three key assumptions for valid causal inference: (1) The genetic variants (instrumental SNPs) are strongly associated with the exposure (relevance assumption), (2) they are independent of confounders of the exposure–outcome relationship (independence assumption), and (3) they affect the outcome exclusively through the exposure, not via any alternative pathways (exclusion restriction assumption or no horizontal pleiotropy).

**(B) Conceptual diagram illustrating bidirectional MR**

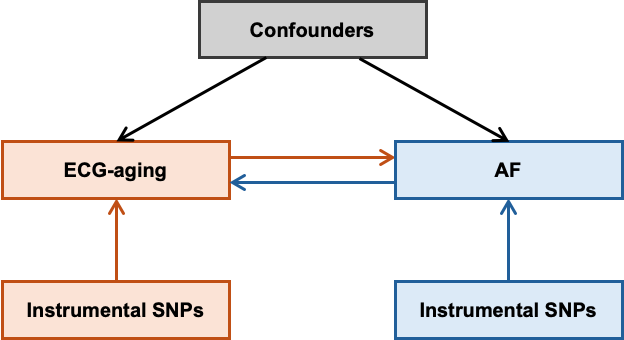

This diagram illustrates the bidirectional MR framework, using distinct genetic instruments to test causal effects in both directions between ECG-aging and AF, while accounting for shared confounders.

**(C) Conceptual diagram illustrating two-step mediation MR**

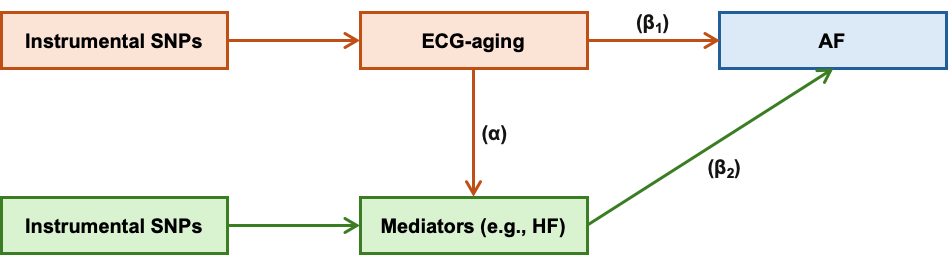

In this two-step MR framework for mediation analysis, the total effect of the exposure (ECG-aging) on the outcome (AF) is partitioned into direct and indirect effects. The indirect effect operates through a mediator (e.g., HF), with α representing the causal effect of ECG-aging on the mediator, and β₂ representing the effect of the mediator on AF, conditional on ECG-aging. β₁ denotes the direct effect of ECG-aging on AF. Distinct genetic instruments are used for the exposure and mediator to minimize horizontal pleiotropy and enable valid estimation of mediation pathways.

Abbreviations: AF, atrial fibrillation; ECG-aging, electrocardiographic aging; HF, heart failure; MR, Mendelian randomization; SNP, single nucleotide polymorphism.

**Supplemental Figure S2. Adjusted cumulative incidence of AF according to AI-ECG age gap quartiles**

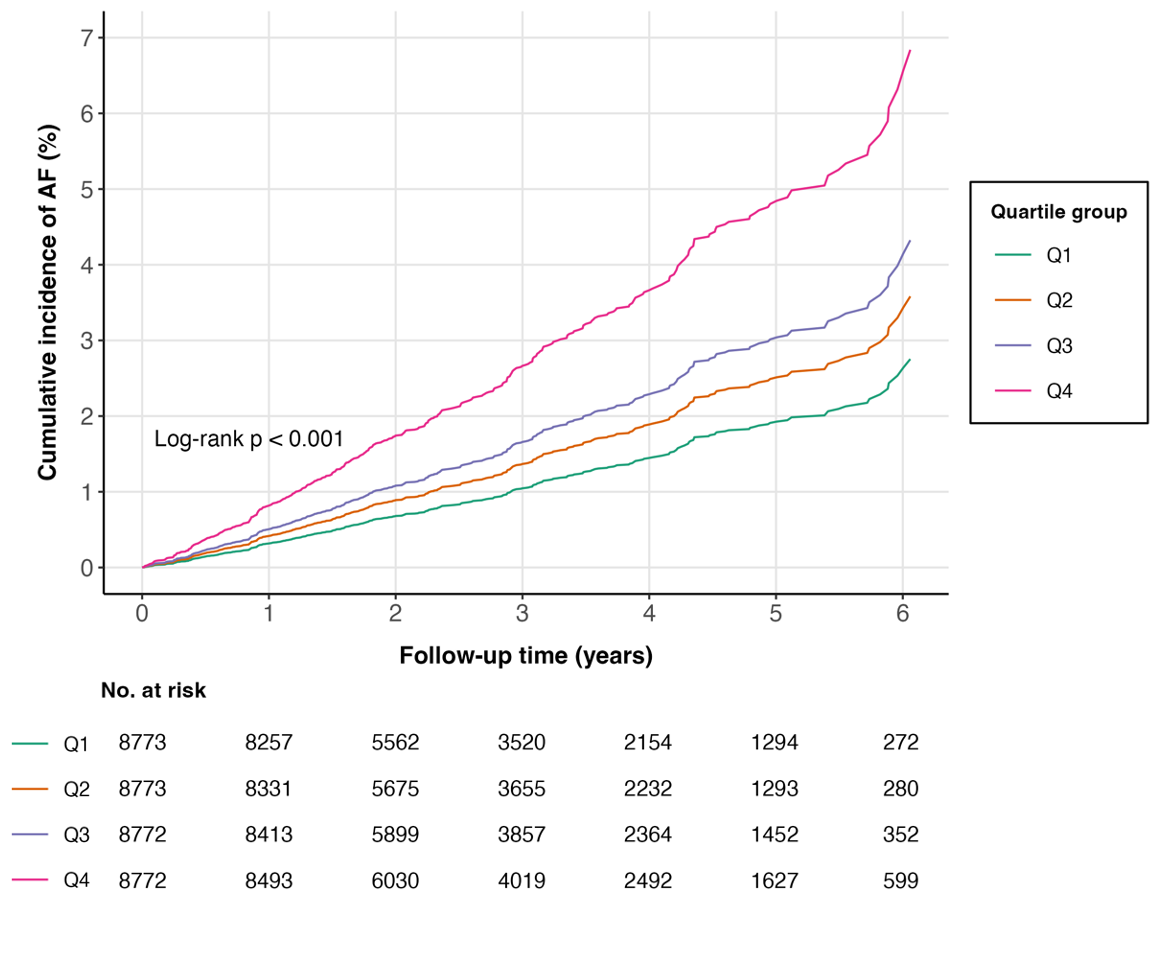

Cumulative incidence curves and corresponding p-values across AI-ECG age gap quartiles were estimated using Cox regression models adjusted for chronological age and sex.

Abbreviations: AF, atrial fibrillation; Q, quartile.

**Supplemental Figure S3. Differences in AI-ECG age gaps between participants with and without prevalent AF at baseline**

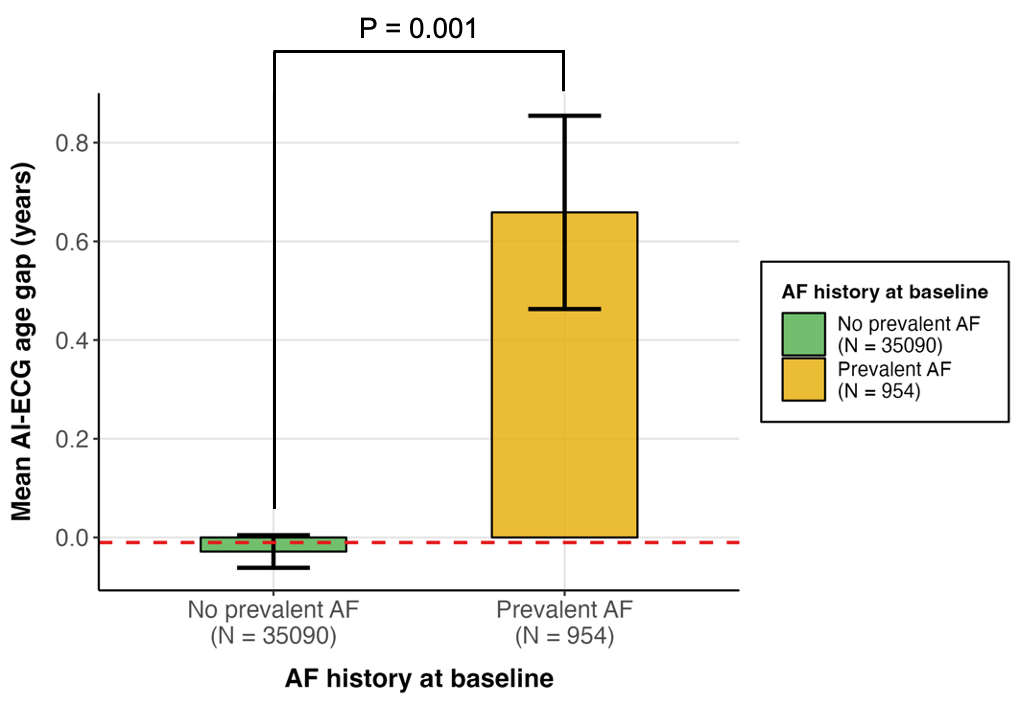

Bar plots depict mean AI-ECG age gaps with error bars indicating standard errors, comparing participants without prevalent AF and those excluded for prevalent AF at baseline. The dashed line represents the overall mean AI-ECG age gap. Group comparisons with corresponding p-values were assessed using the Mann–Whitney U-test.

Abbreviations: AF, atrial fibrillation; AI, artificial intelligence; ECG, electrocardiogram; SE, standard error.

**Supplemental Figure S4. Cumulative incidence curves of AF stratified by GRS percentile groups**

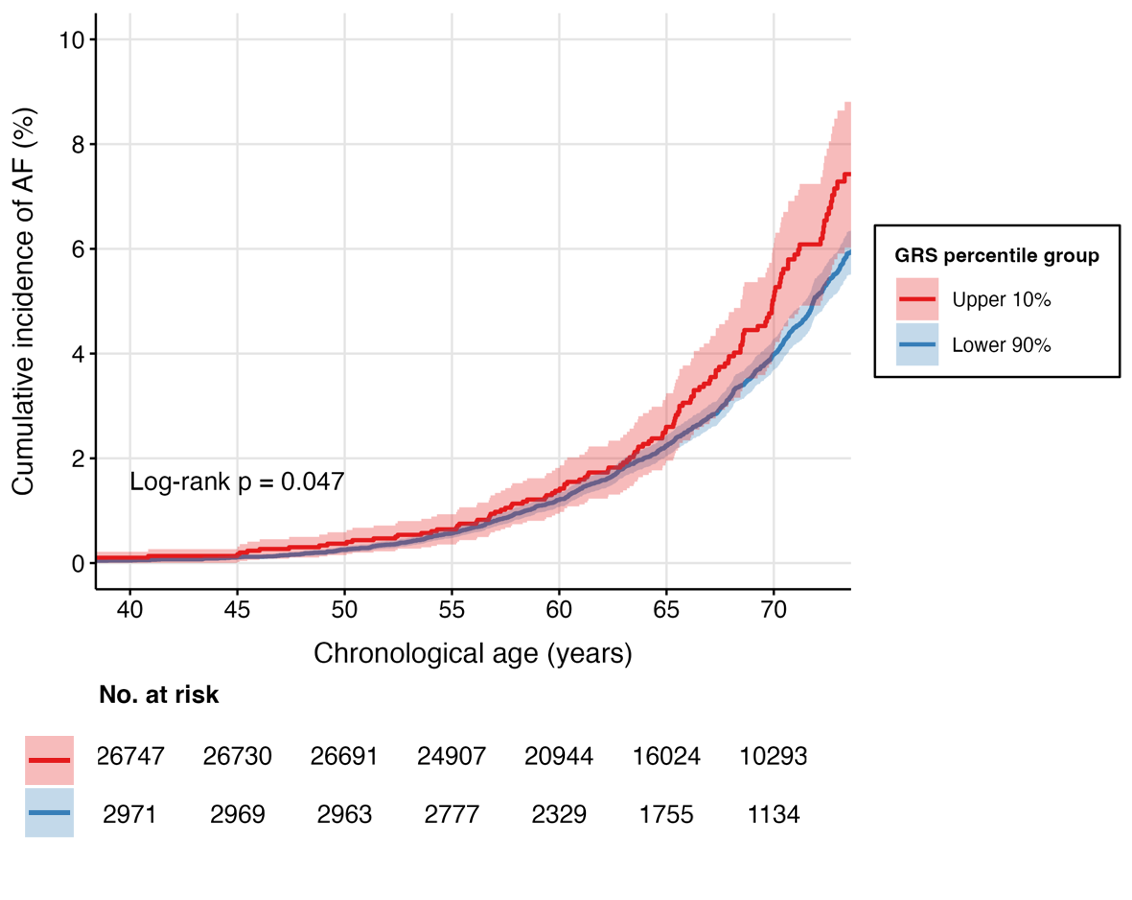

Kaplan–Meier curves illustrate the cumulative incidence of AF between individuals in the top 10% and bottom 90% of the GRS distribution, using chronological age as the time scale. Group comparisons were assessed with the log-rank test, and the shaded areas denote 95% CIs.

Abbreviations: AF, atrial fibrillation; CI, confidence interval; GRS, genetic risk score.

**Supplemental Figure S5. Pyramid plot of observational and genetic associations between AI-ECG age gaps and CMR imaging measurements**

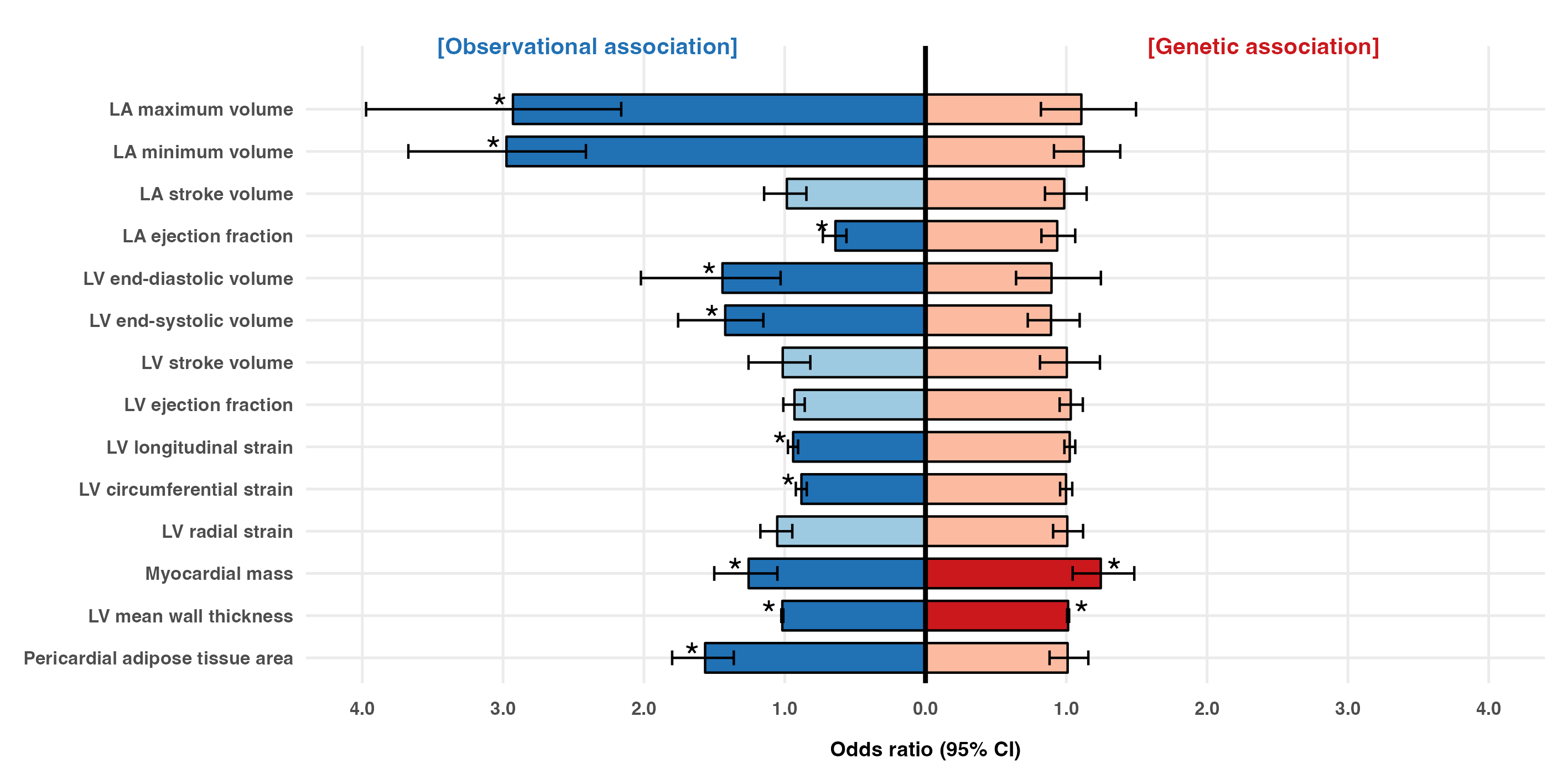

Error bars represent ORs and 95% CIs derived from Model 2 for each CMR imaging parameter, quantified per 1-SD increase in either the measured AI-ECG age gap (left panel) or the GRS for AI-ECG age gap (right panel). Blue and red shading indicate observational and genetic associations, respectively, with darker tones and asterisks marking statistical significance (p < 0.05).

Abbreviations: AI, artificial intelligence; CI, confidence interval; CMR, cardiac magnetic resonance; ECG, electrocardiogram; GRS, genetic risk score; LA, left atrium; LV, left ventricle; OR, odds ratio; SD, standard deviation.

**Supplemental Figure S6. Fractional polynomial Mendelian randomization analysis of the association between genetically predicted ECG-aging and LV remodeling–related CMR traits**

**(A) Myocardial mass (g)**

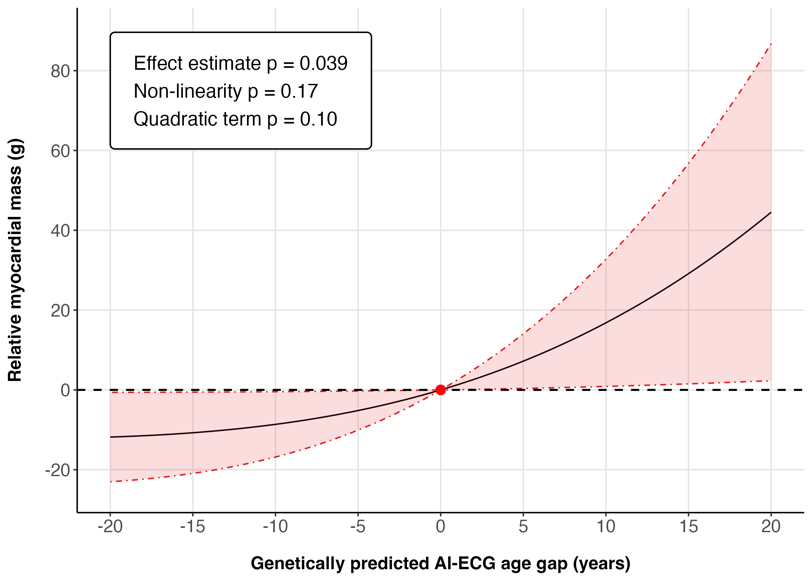

**(B) LV mean wall thickness (mm)**

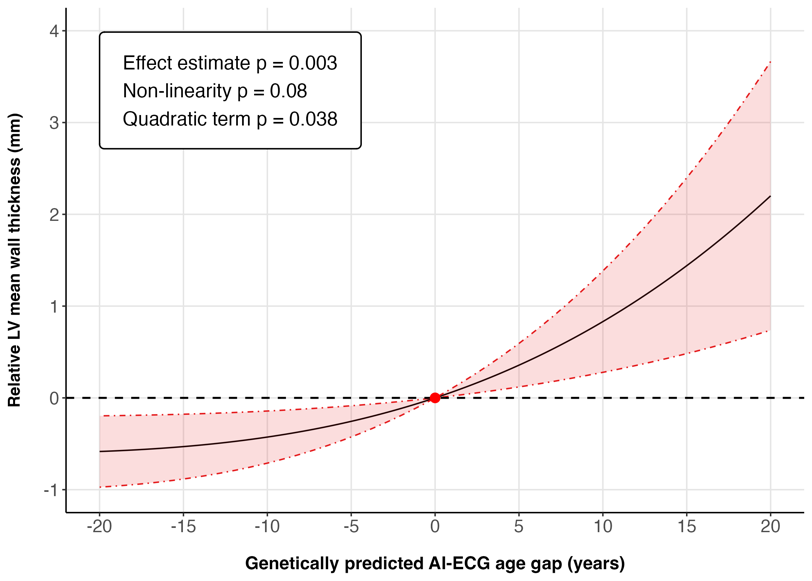

Fractional polynomial MR analysis illustrating the relative beta for each CMR trait across the continuum of genetically predicted AI-ECG age gap. Nonlinear associations were assessed using quadratic and trend tests, with corresponding p-values displayed in each figure. Solid lines denote point estimates, and shaded areas represent 95% CIs.

Abbreviations: AI, artificial intelligence; CI, confidence interval; CMR, cardiac magnetic resonance; ECG, electrocardiogram; ECG-aging, electrocardiographic aging; LV, left ventricle; MR, Mendelian randomization.

**Supplemental Figure S7. Piecewise Mendelian randomization analysis of the association between genetically predicted ECG-aging and LV remodeling–related CMR traits**

**(A) Myocardial mass (g)**

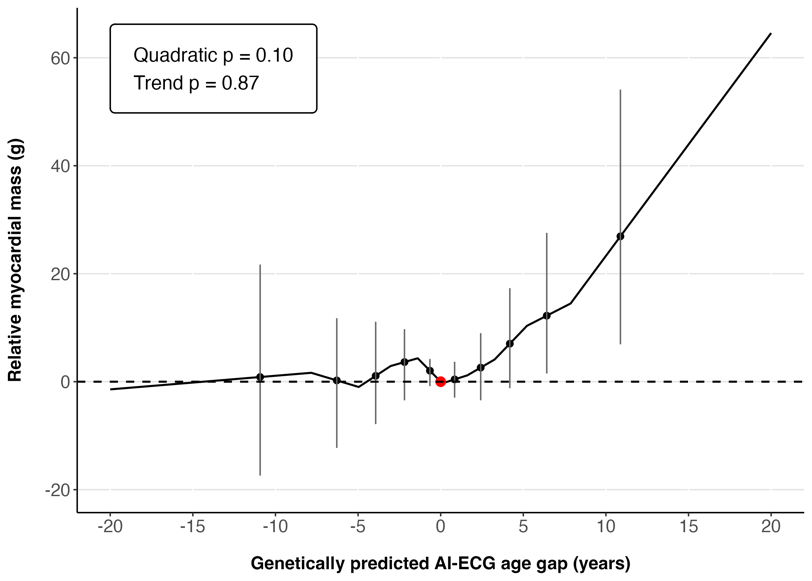

**(B) LV mean wall thickness (mm)**

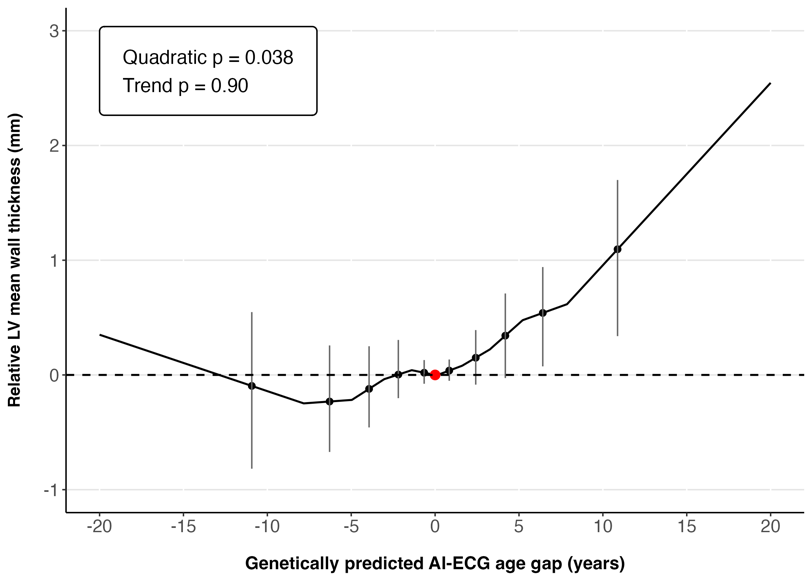

Piecewise MR analysis displaying localized average causal effects across strata of the exposure distribution. The gradient at each value of genetically predicted AI-ECG age gap corresponds directly to the localized average causal effect within that residualized exposure stratum. Nonlinear associations were assessed using quadratic and trend tests, with corresponding p-values displayed in each figure. Solid lines denote point estimates, and vertical error bars represent 95% CIs.

Abbreviations: AI, artificial intelligence; CI, confidence interval; CMR, cardiac magnetic resonance; ECG, electrocardiogram; ECG-aging, electrocardiographic aging; LV, left ventricle; MR, Mendelian randomization.

**Supplemental Figure S8. Scatter plot comparing causal estimates of ECG-aging on AF across multiple Mendelian randomization methods**

**(A) Using GWAS summary statistics for AF from Nielsen et al.**

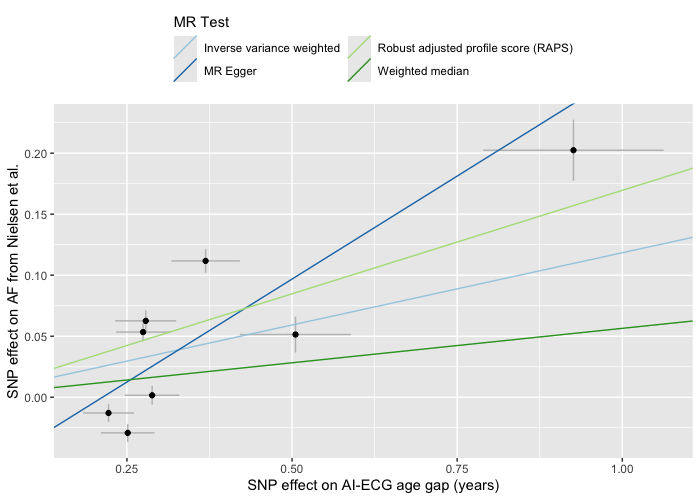

**(B) Using GWAS summary statistics for AF from Nielsen et al. (excluding UK Biobank participants)**

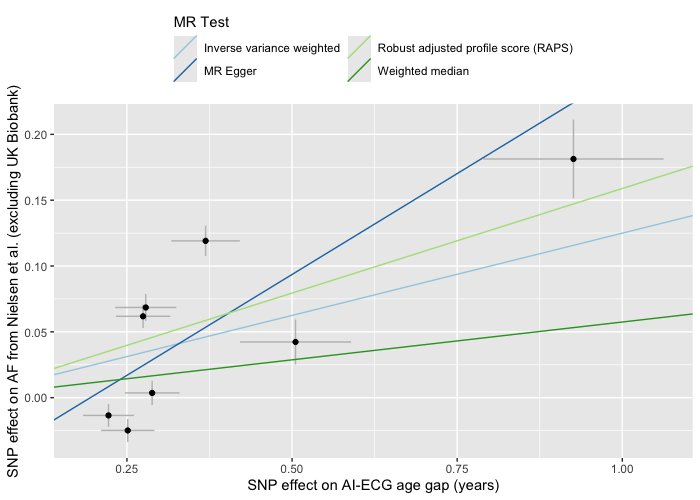

**(C) Using GWAS summary statistics for AF from FinnGen consortium**

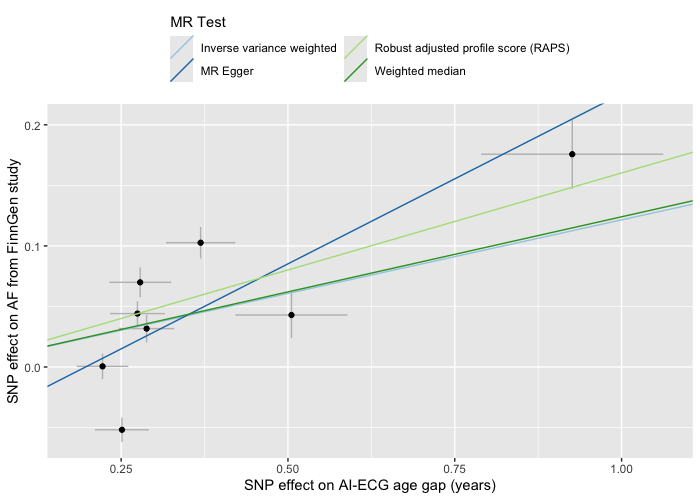

Each panel displays SNP-specific associations between genetically predicted ECG-aging and AF risk paired with regression lines from four MR methods. Dots represent individual SNP effects, with horizontal and vertical error bars corresponding to their standard errors for the exposure and outcome, respectively. Colored regression lines represent the causal effect estimates.

Abbreviations: AF, atrial fibrillation; AI, artificial intelligence; ECG, electrocardiogram; ECG-aging, electrocardiographic aging; GWAS, genome-wide association study; MR, Mendelian randomization; SNP, single-nucleotide polymorphism.

**Supplemental Figure S9. Scatter plot comparing causal estimates of AF on ECG-aging across multiple Mendelian randomization methods**

**(A) Using GWAS summary statistics for AF from Nielsen et al.**

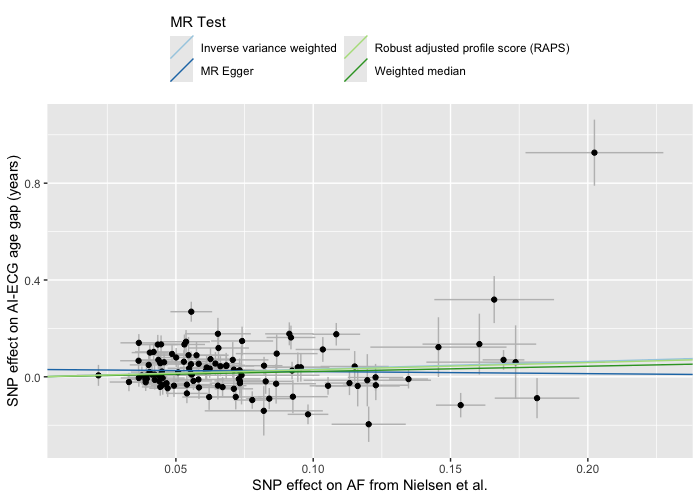

**(B) Using GWAS summary statistics for AF from Nielsen et al. (excluding UK Biobank participants)**

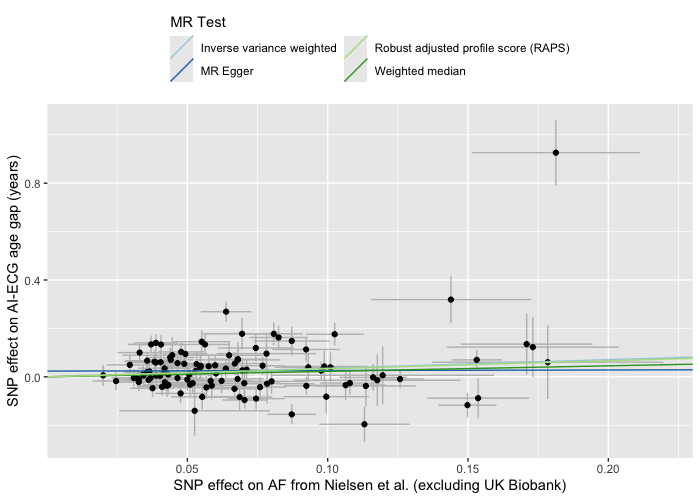

Each panel displays SNP-specific associations between genetically predicted AF risk and ECG-aging paired with regression lines from four MR methods. Dots represent individual SNP effects, with horizontal and vertical error bars corresponding to their standard errors for the exposure and outcome, respectively. Colored regression lines represent the causal effect estimates.

Abbreviations: AF, atrial fibrillation; AI, artificial intelligence; ECG, electrocardiogram; ECG-aging, electrocardiographic aging; GWAS, genome-wide association study; MR, Mendelian randomization; SNP, single-nucleotide polymorphism.

**Supplemental Figure S10. Leave-one-out Mendelian randomization sensitivity analysis for the association between genetically predicted ECG-aging and AF risk**

**(A) Using GWAS summary statistics for AF from Nielsen et al.**

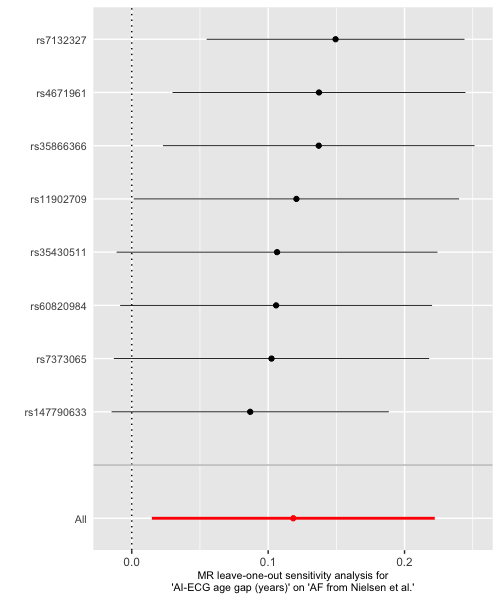

**(B) Using GWAS summary statistics for AF from Nielsen et al. (excluding UK Biobank participants)**

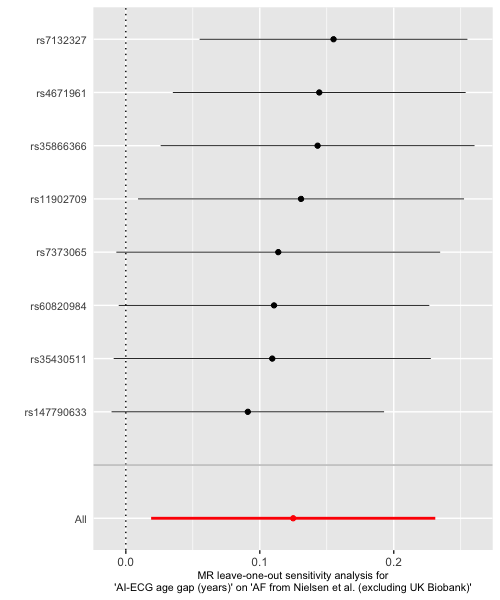

**(C) Using GWAS summary statistics for AF from FinnGen consortium**

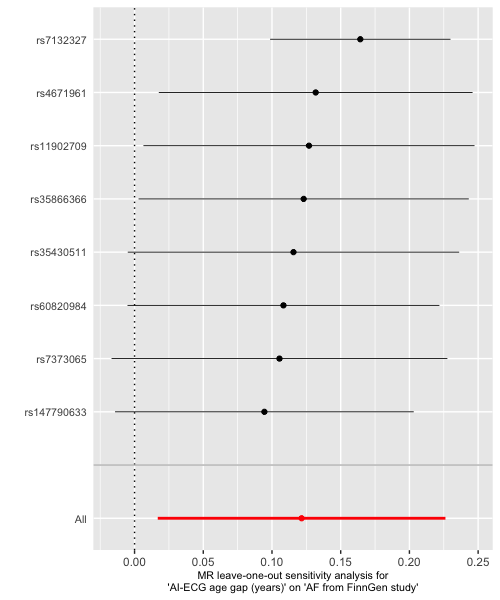

Each panel presents leave-one-out MR estimates, in which the causal effect of genetically predicted ECG-aging on AF risk is recalculated after sequentially excluding each SNP. The bottom red line indicates the overall MR estimate using all genetic instruments.

Abbreviations: AF, atrial fibrillation; AI, artificial intelligence; ECG, electrocardiogram; ECG-aging, electrocardiographic aging; GWAS, genome-wide association study; MR, Mendelian randomization; SNP, single-nucleotide polymorphism.

**Supplemental Figure S11. Leave-one-out Mendelian randomization sensitivity analysis for the association between genetically predicted AF risk and ECG-aging**

**(A) Using GWAS summary statistics for AF from Nielsen et al.**

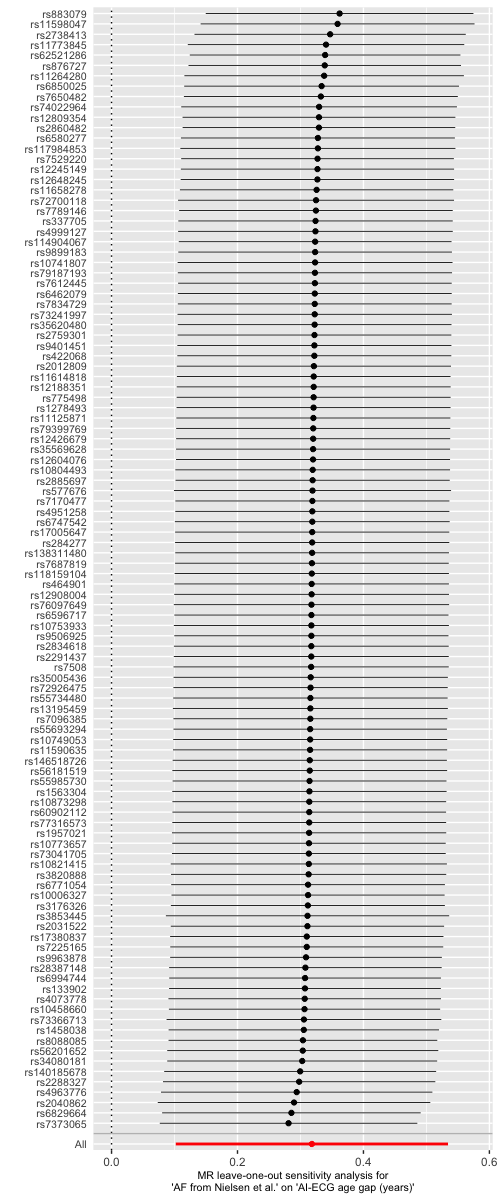

**(B) Using GWAS summary statistics for AF from Nielsen et al. (excluding UK Biobank participants)**

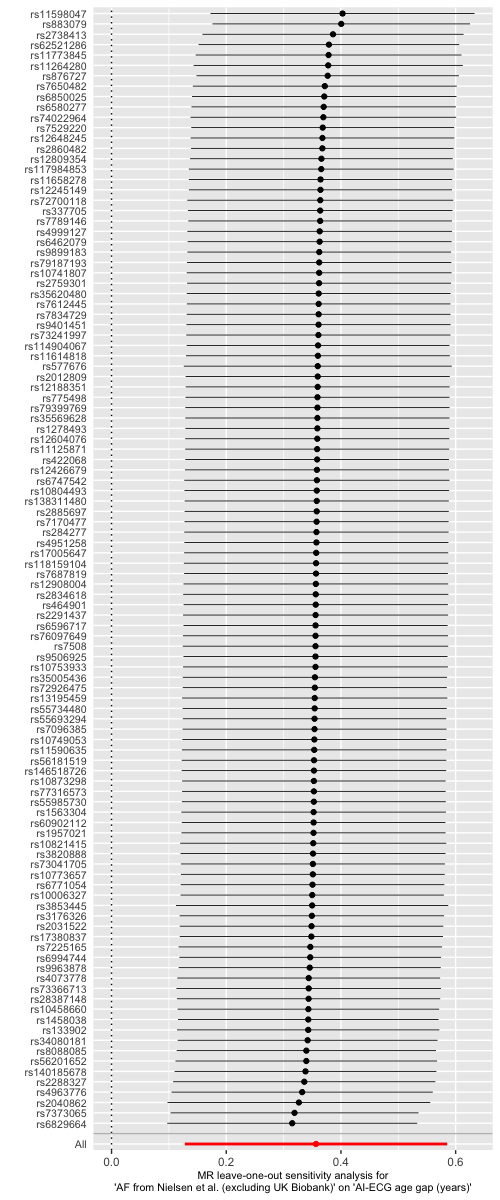

Each panel presents leave-one-out MR estimates, in which the causal effect of genetically predicted AF risk on ECG-aging is recalculated after sequentially excluding each SNP. The bottom red line indicates the overall MR estimate using all genetic instruments.

Abbreviations: AF, atrial fibrillation; AI, artificial intelligence; ECG, electrocardiogram; ECG-aging, electrocardiographic aging; GWAS, genome-wide association study; MR, Mendelian randomization; SNP, single-nucleotide polymorphism.
